# Allosteric DNAzyme for sensitive detection of nucleic acids for molecular diagnosis

**DOI:** 10.1101/2023.08.20.23294196

**Authors:** Chenzhi Shi, Pengfei Wang

## Abstract

Nucleic acids in biofluids are emerging biomarkers for molecular diagnosis of diseases, whose clinical use has been hindered by the lack of sensitive and convenient detection assays. Herein, we report a sensitive nucleic acid detection method based on allosteric DNAzyme biosensors named SPOT (<u>s</u>ensitive loo<u>p</u>-initiated DNAzyme biosens<u>o</u>r for nucleic acid detection) by rationally designing a programmable DNAzyme of endonuclease capability. SPOT can be activated once a nucleic acid target of specific sequence binds to its allosteric module to induce conformational reconfiguration of DNAzyme enabling continuous cleavage of molecular reporters. SPOT provides a highly robust platform for sensitive (LOD: femtomolar for miRNAs, attomolar for SARS-CoV-2 RNA), specific (single-nucleotide discrimination), and convenient (one-step, one-pot, preamplification-free) detection of low-abundant nucleic acid biomarkers. For clinical validation, we demonstrated that SPOT is capable of detecting serum miRNAs (e.g., miR-155, miR-21) from patients for the precise diagnosis of breast cancer, gastric cancer, and prostate cancer. Furthermore, SPOT exhibits potent detection capability over SARS-CoV-2 RNA from clinical swabs with high sensitivity and specificity. Lastly, SPOT is compatible with point-of-care testing modalities such as lateral flow assay to enable convenient visualization. Hence, we envision that SPOT may serve as a robust platform for sensitive detection of a variety of nucleic acid targets towards clinical applications in molecular diagnosis.

## Introduction

Nucleic acids circulating in biological fluids have emerged as prominent liquid biopsy biomarkers for a large diversity of diseases, including cancers and viral infections^1, 2^. For instance, microRNAs (miRNAs) have been revealed to play important roles in many human cancers^3, 4^. Detection of miRNAs in biofluids is therefore a promising strategy for cancer diagnosis, prognosis, and monitoring^5, 6^. While detection of viral nucleic acids such as SARS-CoV-2 RNA is highly essential for infection surveillance and prevention of viral transmission^7, 8^. Nevertheless, a grand challenge ahead of clinical translation of these nucleic acid biomarkers is that they are highly dynamic, heterogenous, and low abundant in biological specimens (e.g., serum, nasopharyngeal swabs)^9-11^. Therefore, there is urgent need for the development of ultrasensitive, specific, and convenient bioassays for the detection of nucleic acid biomarkers possessing significant clinical indications.

Quantitative polymerase chain reaction (qPCR) has been the gold standard laboratory-based analytical technique for sensitive detection of nucleic acids, which, however, requires rigorous sample preparation, expensive instrumentation, and trained personnel to operate in centralized laboratories. Alternatively, numerous isothermal amplification methods with limited reliance on instruments and expertise have been developed for nucleic acid detection, including recombinase polymerase amplification (RPA)^12-14^, loop-mediated isothermal amplification (LAMP)^15-17^, rolling circle amplification (RCA)^18-20^, exponential amplification reaction (EXPAR)^21-23^, hybridization chain reaction (HCR)^24-26^, *etc*. Recently, CRISPR (clustered regularly interspaced short palindromic repeats) technologies have been coupled with isothermal amplifications to realize point-of-care testing (POCT) of a variety of nucleic acids^27-29^, including miRNAs^30, 31^ and SARS-CoV-2 RNA^32, 33^. Although these assays exhibit great sensitivity and simplicity, they may accompany with drawbacks such as non-specific amplifications, serial operational steps, and the need of expensive and delicate enzymes, largely hindering their practical applications^29, 34^. A sensitive and integrated detection assay that spares the need of target amplification and protein-based enzymes, and can be performed in one-pot and in one-step may serve as a good complement to CRISPR-based methods and find its broad application in clinical prospects of nucleic acid biomarker detection-based molecular diagnosis.

DNAzyme is a type of *in vitro* selected, synthetic, all-DNA molecules that exhibits a variety of enzyme-like catalytic activities such as cleaving of nucleic acid phosphodiester bond^35^. A typical nucleic acid-cleaving DNAzyme is composed of a catalytic core (a single-stranded loop with enzymatic capability) flanking with two arms for substrate recognition *via* sequence complementarity. For instance, the classic 10-23 and 8-17 DNAzymes can efficiently cleave ssRNA substrates^36, 37^, while 13PD1 is capable of hydrolyzing ssDNA^38^. DNAzymes have been extensively used for sensing metal ions both *in vitro* and *in vivo* given the fact that its catalytic activity is highly dependent on certain metal ions such as Pb^2+^, Ca^2+^ *etc*^39-42^. Well in contrast, DNAzymes with cofactors like small molecules, proteins, or nucleic acids are difficult to select thus rarely reported, making it hard to achieve direct detection of such targets^35, 43^. Instead, inspired from natural enzymes and pioneering works of ribozymes^44, 45^, allosteric DNAzyme biosensors have been designed by implementing an allosteric module (e.g., aptamer, toehold) to mediate its catalytic activity via modulators such as small molecules^46-48^, proteins^49-52^, nucleic acids^53-61^, or bacteria^62, 63^.

Conventional allosteric DNAzyme biosensors for nucleic acid targets are generally designed into a multicomponent molecular complex (Fig. S1), either by inhibiting and freeing the DNAzyme *via* an inhibitor strand of toehold^53^, or by splitting and resuming the catalytic core upon target binding^58, 64, 65^, or by target-induced stabilization of DNAzyme-substrate-target complex^66^. These biosensing systems typically exhibit a picomolar to nanomolar sensitivity for direct detection against targeting nucleic acids^58^, which largely fall short in meeting the need of clinical detection of miRNAs or viral RNA biomarkers. We propose that certain drawbacks may accompany with these systems due to constrains existing in the nature of multicomponent designs that negatively impact its detection capability for low-abundant targets. First, the multicomponent DNAzyme systems (i.e., the toehold-mediated system) demand precise stoichiometry between composing strands for sensing trace amount of target, which are extremely hard to achieve in practice. Second, molecular complexes of multiple strands are vulnerable to unwanted molecular interaction-caused kinetic traps thus requires comprehensive optimization of sequence designs. Third, the splitting-and-resuming of DNAzyme from separate strands may impede its catalytic activity^64^. Fourth, in these designs, though thermodynamically unfavorable, the substrate is able to interact with the DNAzyme due to partial exposure of its flanking arm, which may induce leakage of signal. One earlier work integrates a molecular beacon and DNAzyme into a single strand to allosterically mediate the catalytic activity of DNAzyme *via* target binding-induced stem opening, which, however, only realizes a nanomolar detection sensitivity, presumably due to high background signal derived from partial exposure of the binding arm^54^. Herein, to address these above challenges, we engineer the allosteric DNAzyme into a one-stranded, self-locked molecular system composing of an intact catalytic core and fully sealed flanking arms, with an internal loop serving as the allosteric module for nucleic acid detection (Fig. 1), named SPOT (<u>s</u>ensitive loo<u>p</u>-mediated allosteric DNAzyme biosens<u>o</u>r for nucleic acid detection). We conducted comprehensive computational and experimental studies to pinpoint the optimal designs of SPOT with high detection capability, and fully revealed its molecular mechanism for DNAzyme activation upon target hybridization. The SPOT assay exhibited significantly enhanced detection sensitivity (15 fM for miR-21, 1.9 aM for viral RNA) and specificity (single-nucleotide discrimination) on nucleic acid targets by using a one-pot, one-step, preamplification-free, and isothermal detection protocol. To validate its clinical application, we demonstrated that SPOT can realize highly sensitive and specific detection of serum miRNAs for the precise molecular diagnosis of multiple cancers including breast cancer, gastric cancer, and prostate cancer. In addition, SPOT is capable of probing SARS-CoV-2 RNA from clinical swabs with high sensitivity and accuracy. Lastly, SPOT is readily adaptable to couple with POCT modalities such as lateral flow assay (LFA) to achieve instrument-free detection of nucleic acid targets. We envision that SPOT shall find its broad applications in clinical prospects especially in liquid biopsy-based detection of circulating single-stranded nucleic acid biomarkers towards molecular diagnosis of various diseases.

**Fig. 1.**
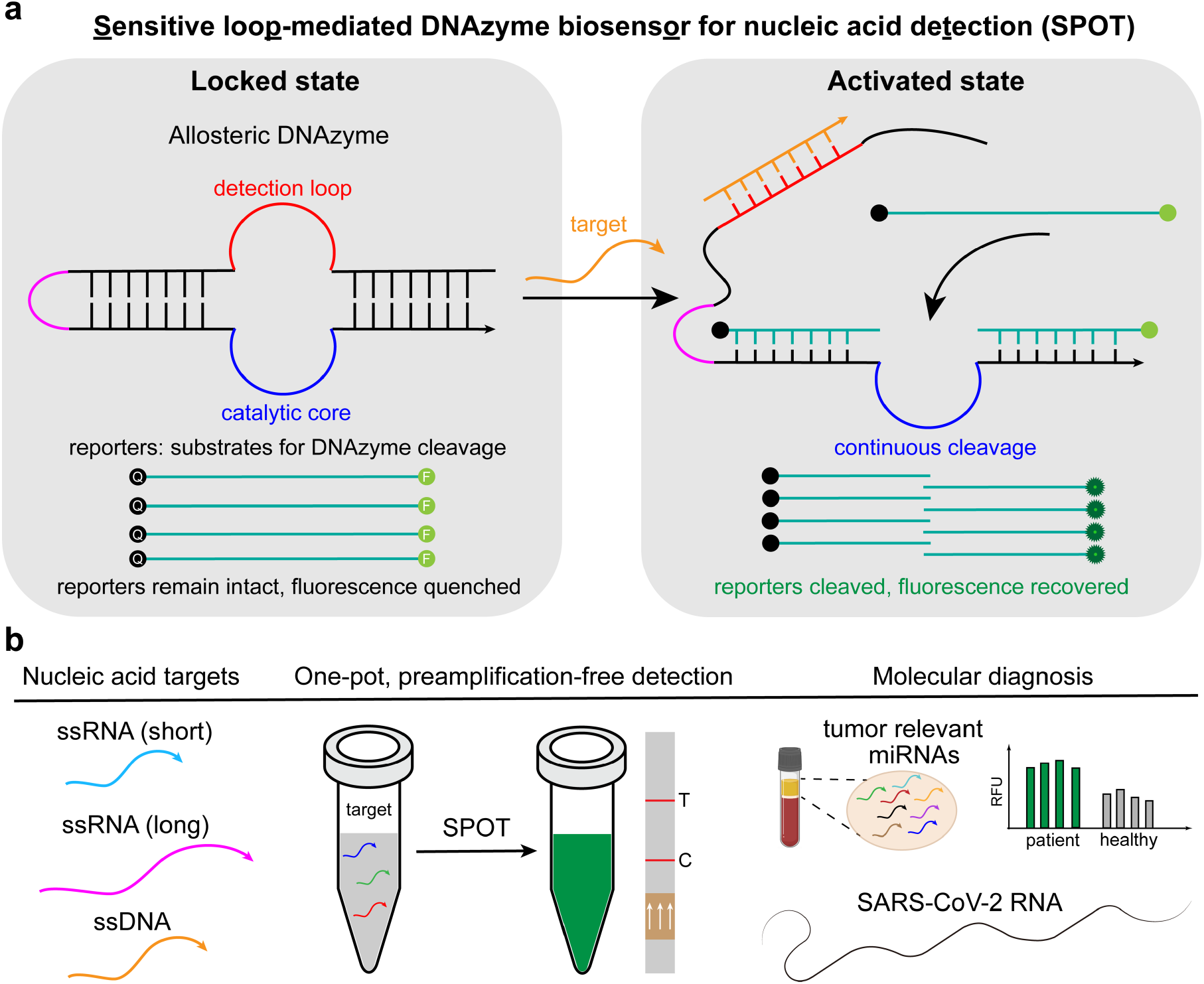
Overview of SPOT. **a**, Working principle of SPOT. DNAzyme of endonuclease activity (10-23 DNAzyme is used for most of the designs) is locked by extending its 5’ end for hybridizing and blocking the two flanking arms to disable its substrate-binding capability without interfering the catalytic core. A single-stranded loop is integrated between the two locking domains serving as the allosteric module for cofactor recognition and detection. SPOT remains inactive in its locked conformation as the flanking arms are fully sealed. Once a ssRNA or ssDNA target binds to the detection loop, the as formed double-stranded duplex introduces mechanical tension between the two locking domains that renders them thermodynamically unstable thus leads to the opening of the DNAzyme for continuous substrate cleavage for signal generation. **b**, One-pot, one-step, preamplification-free, and isothermal detection of various single stranded nucleic acid targets by SPOT for molecular diagnosis of diseases including cancer and SARS-Cov-2 infection.

## Results

### Working principle of SPOT

SPOT is designed by elongating the 5’ end of the DNAzyme to self-fold and hybridize with the two flanking arms forming locking domains to prevent substrate from binding (Fig. 1a). A single-stranded loop of specific sequence is placed in between the locking domains serving as the allosteric module to modulate the catalytic activity of DNAzyme. Once the allosteric cofactor, single-stranded nucleic acids in this case, binds to the loop *via* sequence complementarity, the as formed double-stranded rigid DNA duplex introduces mechanical tension between the two locking domains which subsequently leads to partially or fully exposure of the flanking arms, similar to the working mechanism of molecular beacons^67^. The nucleic acid substrates then continuously bind to and get cleaved by the DNAzyme for fluorescent signal amplification, whose fluorescence are initially quenched by an adjacent quencher due to proximity. SPOT exhibits several distinct characteristics comparing to existing DNAzyme-based biosensing systems. Firstly, SPOT is built from a single intact DNA strand with both flanking arms fully sealed, which not only maintains the high cleaving activity of DNAzyme (leads to higher signal), realizes better assembling and locking efficiency given intramolecular interaction is thermodynamically more favorable and stable (leads to lower noise), but also avoids stringent requirement over stoichiometry precision of various DNA strands as in multicomponent molecular systems (ease of operation). Secondly, SPOT can be modularly designed and rationally optimized by programming the sequences of the detection loop and flanking arms against multiplex distinct targets of high specificity and orthogonality. Thirdly, SPOT holds high detection capability enabling it capable of integrating the whole assay into one-step and one-pot by sparing the need of target preamplifications. While at the meantime, comparing to CRISPR-based diagnostic assays, SPOT provides a relatively simple, robust, and cost-effective platform since no protein-based expensive and delicate enzymes are used. Such a vastly simplified assay negates the need for sophisticated instrumentations, which offers an adaptable modality for POCT of nucleic acid biomarkers for the molecular diagnosis of a variety of diseases including cancer and viral infections (Fig. 1b).

### Design and screening of SPOT for nucleic acid detection

10-23 DNAzyme is reported having the highest catalytic activities for cleaving single-stranded RNA substrates in a sequence-specific manner^35, 36^, which therefore has been chosen for SPOT construction. Prior to designing and screening SPOT, we examined the cleaving capability of the free-form 10-23 DNAzyme on four types of substrates (Fig. 2a), including a linear all-RNA (R-L) reporter, a linear DNA-RNA chimera (D/R-L) reporter, two DNA-RNA chimera molecular beacon (MB) reporters with 3-bp-long or 4-bp-long stems (D/R-MB3, D/R-MB4). The DNAzyme is flanked with two 7-nt-long binding arms, which was reported to have one of the fastest cleaving rate^68^. As revealed, 10-23 DNAzyme showed potent cleaving activity against all four reporters (Fig. 2b), with D/R-L and D/R-MB4 reporters having the highest signal-to-noise ratio (SNR) after 120 mins of reaction (Fig. 2c, Fig. S2). D/R-L reporter was selected for later experiments given its design simplicity.

**Fig. 2.**
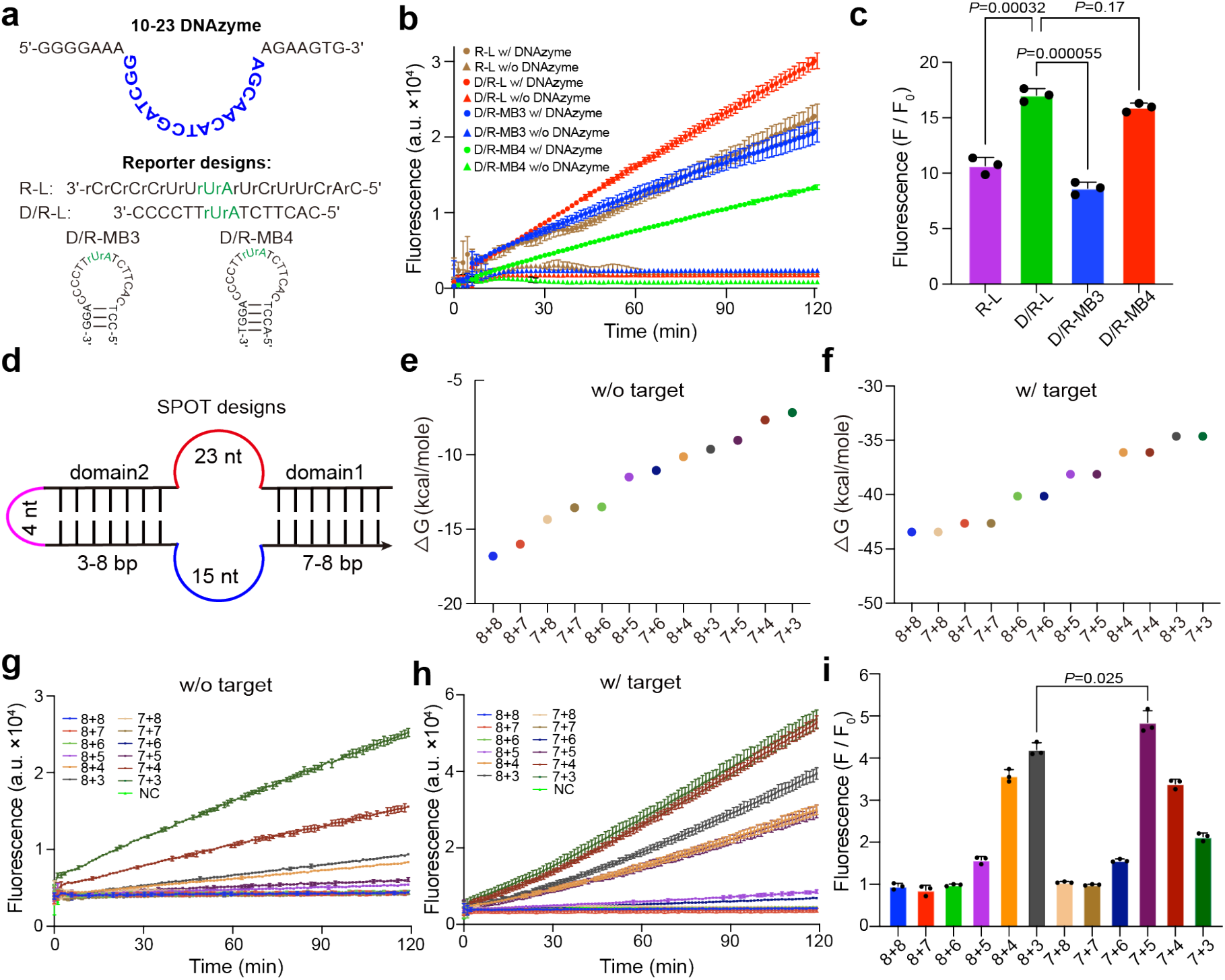
Design and screening of SPOT constructed from 10-23 DNAzyme. **a**, Design of free-form 10-23 DNAzyme for RNA or DNA-RNA chimera substrate cleavage. **b**, Cleavage of various substrates over a time course of 120 mins. **c**, Fluorescence ratio (F/F_0_) of signal (F, with DNAzyme) to background noise (F_0_, without DNAzyme) after 2 hours of cleavage against various reporters. **d**, Schematic designs of SPOT based on 10-23 DNAzyme. **e**, Calculated Gibbs free energy change for SPOTs in the absence of target nucleic acids. **f**, Calculated Gibbs free energy change for SPOTs with target nucleic acids binding to the detection loop. **g**, Fluorescence change of reporters (F_0_) over a time course of 120 mins while incubating with SPOTs without activation by target nucleic acids. SPOT: 10 nM, reporter: 500 nM. **h**, Fluorescence change of reporters (F) over a time course of 120 mins while incubating with SPOTs that have been activated by target nucleic acids. SPOT: 10 nM, miR-155: 10 nM, reporter: 500 nM. **i**, Fluorescence signal-to-noise ratio (SNR) of SPOTs for the detection of miR-155 after 120 mins of incubation. SPOT-7+5 design exhibits the highest SNR. All experimental measurements are mean ± standard deviation (SD) with n = 3. *P* values were calculated by two-tailed Student’s t-test.

There are four aspects need to be taken into account while designing SPOT to achieve high SNR for target detection. First, the length of locking domains. Longer domains have stronger locking effect to minimize leakage, but it may hinder the opening of SPOT after target binding. Second, the length of detection loop. Longer loop promotes the opening of SPOT given the formation of longer rigid duplex after target binding, but at the meantime it may induce higher instability thus leakage to SPOT attributed from increased chance of unwanted molecular interactions. Third, the length of RNA substrates. An appropriate length of RNA substrate is needed in order to achieve fast on-off rate for efficient cleavage. Fourth, the length of the linker segment. Longer linker may cause miss interactions, while a too short linker may impede self-folding of locking domains due to limited molecular flexibility. Of note, it is impossible to realize an optimal design for one aspect without interfering others, instead, these parameters need to be leveraged to pinpoint the SPOT design with high SNR for target detection. To simplify the design and screening of SPOT, the detection loop is initially set as 23-nt-long that is complementary to a miRNA target (miR-155), which is a cancer relevant biomarker^69^. Furthermore, we set the RNA substrate as 15-nt-long, with two 7-nt segments respectively binding to the two flanking arms of the DNAzyme, as demonstrated above. In addition, the linker is fixed at 4-nt long, which shall provide appropriate flexibility without causing apparent molecular interference. With these parameters being fixed, locking domains of various lengths are then designed for screening (Fig. 2d). The rationale for designing locking domains is to make it sufficiently strong to prevent leakage, while not too strong to hamper opening after target binding. With this rationale in mind, the length of domain1 is set at 7-bp or 8-bp, and the length of domain2 is set at 3-bp to 8-bp (Fig. S3).

Computational studies were first conducted to theoretically investigate SPOT designs. The secondary structures of SPOTs were simulated and their Gibbs free energy change were calculated by using NUPACK^70^. As illustrated (Fig. S4), most SPOTs showed expected molecular structures with both locking domains fully sealed, with the exception of SPOT-7+3, SPOT-7+4, SPOT-8+3, and SPOT-8+4, whose domain2 cannot be locked due to interactions between the linker and the detection loop, suggesting these SPOTs may have severe leakage issue. For other designs, molecular interactions of 4-bp binding between the detection loop and the catalytic core may be observed, which shall have neglectable effect on target binding given such weak interactions. Calculated Gibbs free energy change was plotted in Fig. 2e, validating that SPOTs of longer locking domains are thermodynamically more stable. After addition of target, simulated secondary structures of SPOT-target complexes suggested the successful binding of target to the detection loop leading to opening of locking domains for substrate invasion (Fig. S5), whose Gibbs free energy change was illustrated in Fig. 2f.

We next sought to experimentally study the detection performance of SPOTs with miR-155 as the targeting nucleic acid. Prior to comprehensive screening, we investigated the experimental parameters of ionic strength, temperature, DNAzyme concentrations *etc*. and have identified the relatively optimal conditions for SPOT (Fig. S6). Screening results revealed that most SPOTs showed little cleavage of reporters thus minimal leakage prior to adding miR-155, while SPOT-7+3, SPOT-7+4, SPOT-8+3, and SPOT-8+4 exhibited significant signal leakage (Fig. 2g), agreeing well to computational predictions (Fig. S4). Upon target binding to the detection loop, apparently elevated fluorescence over time was observed for SPOT-7+3, SPOT-7+4, SPOT-8+3, SPOT-8+4, and SPOT-7+5, but the other designs showed minimal reporter cleavage (Fig. 2h), suggesting they cannot be activated. The trend of cleavage activity is in good agreement to the thermodynamic stability of SPOTs while complexing with targets (Fig. 2f). The experimental results agreed well to the thermodynamic calculations and to our speculations that SPOTs of long locking domains are hard to be activated in the presence of targets, while SPOTs of short locking domains are associated with signal leakage in the absence of targets. Amongst SPOTs being examined, SPOT-7+5 exhibits the highest SNR for detecting miR-155 targets (Fig. 2i), which was therefore selected for subsequent detection experiments unless otherwise specified. We then used native polyacrylamide gel electrophoresis (PAGE) to further characterize the assembly and activation of SPOT-7+5 (Fig. S7). It was revealed that the target miR-155 can readily bind to SPOT forming a larger complex of retarded motility, which induced significant cleavage of the RNA reporters comparing to inactivated SPOT, validating the successful construction of SPOT for nucleic acid detection.

To explore the design generality of SPOT beyond 10-23 DNAzyme, we employed a DNA-hydrolyzing 13PD1 DNAzyme for SPOT construction, which is composed of a catalytic core of 41-nt flanked by two binding domains.^38^ Free-form 13PD1 showed apparent cleavage of DNA substrates only at a relatively high concentration of DNAzyme (Fig. S8), suggesting the limited catalytic activity of 13PD1 comparing to 10-23 DNAzyme. SPOTs based on 13PD1 were designed, screened, and then selected for target nucleic acid (a DNA mimic of miR-155) detection (Fig. S9). It was revealed that SPOT of 13PD1 can be readily activated by the single-stranded DNA target, supporting that this design strategy may be well translated to other types of DNAzymes beyond 10-23.

### Mechanistic studies of SPOT

We then conducted mechanistic studies to thoroughly unveil the molecular mechanism of SPOT for target detection, specifically the 10-23 DNAzyme SPOT with 7+5 bp locking domains and a 23-nt detection loop. As predicted by NUPACK simulations, binding of target to detection loop induces opening of domain1, while with domain2 remaining locked (Fig. 3a, Fig. S5). To further validate this computational prediction, we used oxDNA to conduct molecular dynamics (MD) simulation of SPOT by taking into account of both thermodynamic and mechanical properties of DNA strands.^71^ MD simulation of SPOT also reveals that the binding of target leads to the opening of domain1 but not domain2 (Fig. 3b), in good agreement to NUPACK simulations. Changes of distances between selected bases in MD models within locking domains prior to (Fig. 3c, Fig. S10) and after (Fig. 3d) addition of target further validated their molecular configurations.

**Fig. 3.**
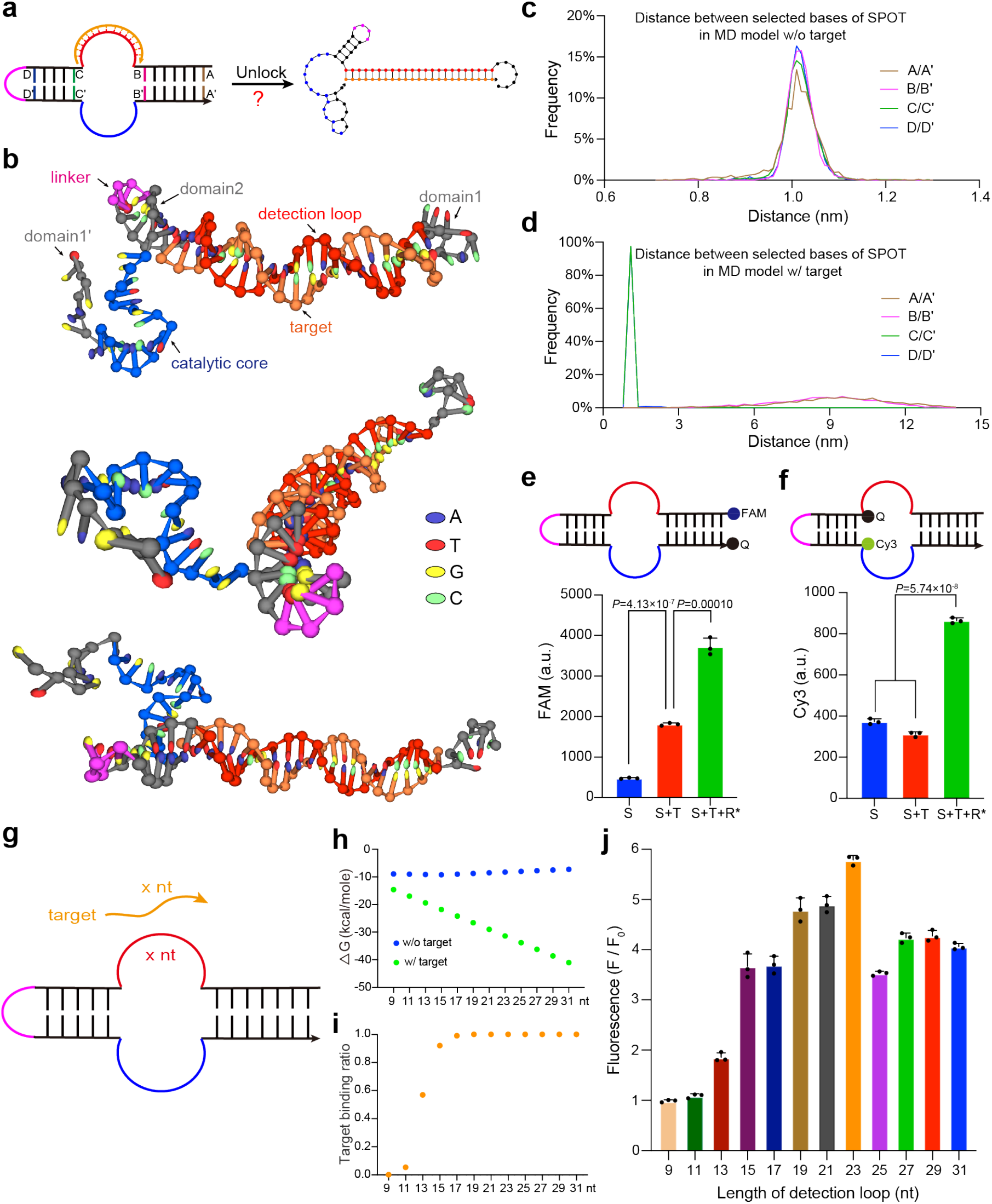
Mechanistic studies of SPOT. **a**, Molecular activation pathway of SPOT upon target binding to the detection loop as suggested by NUPACK simulations. **b**, oxDNA simulation of SPOT-7+5 after target binding. The molecular conformation is presented from three different views. **c, d**, Distances between designated bases (as indicated in **a**) in the MD models of SPOT-7+5 prior to (**c**) and after (**d**) addition of target. **e, f**, FRET experiments to investigate the molecular conformational change of SPOT-7+5 after target hybridization. S: SPOT; T: miR-155 target; R*: DNA mimic of the RNA reporter to avoid being cleaved by SPOT thus maintain it in its open state. **g**, Designs of SPOT-7+5 with various detection loop lengths. **h**, Calculated Gibbs free energy change for SPOT-7+5 of various detection loops without or with target binding. **i**, Binding ratio between targets and SPOTs of various detection loops. **j**, Fluorescence SNR of SPOTs of various detection loops for the detection of corresponding RNA targets. F: fluorescence in the presence of target. F_0_: fluorescence in the absence of target. All experimental measurements are mean ± SD with n = 3. *P* values were calculated by two-tailed Student’s t-test.

To experimentally verify the molecular activation pathway of SPOT, fluorophores are respectively anchored at the ends of two locking domains, whose fluorescence are initially quenched by adjacent quenchers due to proximity when SPOT is in locked conformation and then recovered if that locking domain gets opened after target binding (Fig. 3e,3f). As revealed, after adding miR-155 to SPOT, a significant increase in fluorescence was observed for FAM but not for Cy3, indicating that domain1 was opened while domain2 remained locked. Addition of the uncleavable DNA substrates led to significantly enhanced fluorescence recover for both fluorophores due to further enlarged distance between fluorophores and quenchers under fully unlocked conformation. Taken together, both computational and experimental studies revealed the molecular unlocking mechanism of SPOT: domain1 is opened upon target hybridization, which leads to subsequent substrate invasion, strand displacement-induced opening of domain2, and finally cleavage of substrates (Fig. S11). Of note that the successful opening of domain1 by target hybridization may not necessarily enable substrate binding and cleavage given the fact that domain2 remains locked for SPOT designs of longer domains, meaning that the substrate must be capable of unlocking domain2 by itself. For instance, though domain1 of SPOT-8+8 can be opened after introducing the target, no apparent substrate cleavage was observed given the substrate was incapable of effectively opening domain2 (Fig. 2h, Fig. S5, Fig. S12).

We next investigated how the length of detection loop may affect the performance of SPOTs. SPOTs (7+5-bp) with detection loops ranging from 9 to 31-nt were designed for examination (Fig. 3g). To minimize the confounding effect of sequences, the detection loop is composed of single-stranded polythymine for the detection of polyadenine targets of the same length. Without targets, NUPACK simulations predicted the formation of SPOTs of similar molecular structures and calculated Gibbs free energy changes (Fig. 3h, Fig. S13). After target hybridization to the detection loop, as expected, SPOTs of longer loops showed larger changes in Gibbs free energy, thus thermodynamically more favorable to form and to activate DNAzyme (Fig. 3h, Fig. S14). Noted that SPOTs with detection loops of 9-nt, 11-nt, and 13-nt long showed very low target-detection loop binding ratio, suggesting they are difficult to be activated (Fig. 3i). We then conducted experimental examinations of SPOTs with various detection loop lengths. In consistent to above simulations, SPOTs with detection loops of 9 to 13-nt-long yielded the lowest SNR that close to one because the targets were unable to bind to their detection loops (Fig. 3j). Efficient activation of SPOT begins once detection loop is increased to 15-nt, with 19-nt, 21-nt, and 23-nt designs exhibiting the highest SNRs. Though longer detection loop (e.g., 25 to 31-nt designs) leads to stronger SOPT activation upon target recognition, but it also accompanies with higher leakage (Fig. S15), thus rendering decreased SNR. Similar trend may also be observed for the detection of elongated and truncated miR-155 targets of various lengths (Fig. S16).

### Examining the detection capability of SPOT

Having constructed SPOT and clarified its working mechanism, we next examined SPOT’s detection capability on various miRNA targets (Fig. 4, Fig. S17). The detection sensitivity of SPOT was investigated by using three miRNA targets including miR-155, miR-21, and miR-16. miRNA targets of a series of concentrations ranging from 0 to 50 pM were subject to SPOT detection, as expected, whose fluorescence values are positively related to miRNA concentrations (Fig. 4a-c). A statistical significance in detection could be observed for as low as 100 fM for all three miRNA targets. A linear relationship between fluorescence and logarithm of miRNA concentrations was revealed in Fig. 4d-f, with a calculated LOD of 19 fM, 15 fM, and 91 fM for miR-155, miR-21, miR-16, respectively. These experiments demonstrated that SPOT shall be sufficiently sensitive for detecting biological circulating miRNA targets given that their abundance was reported to most likely fall within the picomolar range^72^. Specificity represents another essential property for a detection assay. We thoroughly characterized the detection specificity of SPOT by using miR-155 as the targeting miRNA. As revealed,

**Fig. 4.**
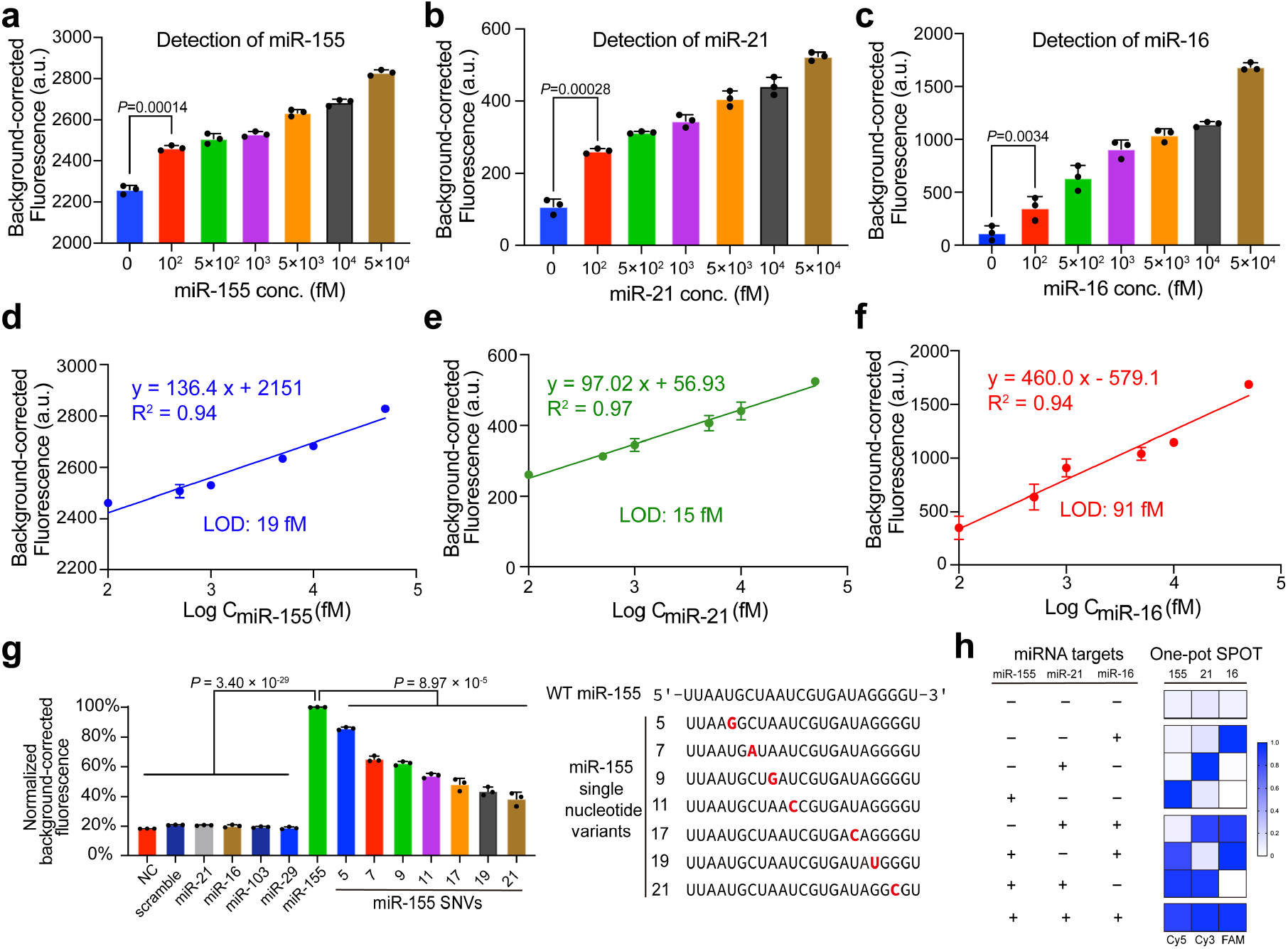
Detection capability of SPOT for synthetic miRNA targets. **a**-**c**, Background-corrected fluorescence generated by SPOT after incubating with different concentrations of miR-155, miR-21, and miR-16, respectively. SPOT: 10 nM; reporter: 500 nM; incubation time: 2 hr. **d-f**, Corresponding linear curve between fluorescence and logarithm of miRNA concentrations. LOD of 19 fM, 15 fM, and 91 fM are calculated based on this curve for the detection of miR-155, miR-21, and miR-16, respectively. **g**, Examination of detection specificity of SPOT targeting miR-155. miRNAs of distinctly different sequences or single nucleotide variants of miR-155 were detected and compared to the wild type miR-155 target. Background-corrected fluorescence was normalized by setting miR-155 as 100%. SPOT: 10 nM; target: 1 nM; reporter: 500 nM; incubation time: 2 hr. **h**, Investigating the orthogonality of SPOT for detecting multiple miRNA targets in a one-pot assay. Fluorophores: Cy5 for miR-155, Cy3 for miR-21, FAM for miR-16. SPOT: 10 nM; target: 10 nM; reporter: 500 nM; incubation time: 2 hr. All measurements are mean ± SD with n = 3. *P* values were calculated by two-tailed Student’s t-test.

RNA targets of distinct sequences different to miR-155 led to neglectable and insignificant fluorescence increase of SPOT as comparing to the miR-155 target (Fig. 4g). Single nucleotide mutations were incorporated at various positions of miR-155 to examine whether SPOT is able to discriminate single nucleotide variants (SNV) from wild type (WT) miR-155. Comparing to WT, miR-155 SNVs exhibited significantly lower fluorescence, indicating that SPOT is capable of single nucleotide discrimination. Interestingly, SPOT showed better discrimination for SNVs composing mutations closer to the 3’ end, which we believe is in perfect agreement to the molecular unlocking pathways where mutations closer to the 3’ end rendering the target less capable of opening locking domain1 for SPOT activation.

One of the advantageous features of SPOT is its reporter cleavage specificity since DNAzyme cleaves its substrates in a highly sequence-specific manner, unlike the nonspecific *trans*-cleavage of reporters by Cas endonucleases. This cleavage specificity enables SPOT of high design orthogonality and multiplexity in a one-pot detection systems for various different nucleic acid targets. For demonstration, we designed three different SPOT systems of unique detection loops and fluorescent reporters against miRNA targets of miR-155 (Cy5), miR-21 (Cy3), and miR-16 (FAM), respectively (Fig. S17). In a one-pot multiplex system, SPOT selectively responded to its target miRNAs with little cross-talk observed (Fig. 4h), proving the high orthogonality and multiplexing capability of SPOT for miRNA detection.

### Detection of serum miRNAs for cancer diagnosis

As a proof-of-concept demonstration for clinical applications, we utilized SPOT for the detection of serum miRNAs for diagnosis of multiple types of cancers (Fig. 5). Total miRNAs were firstly extracted from serum and then subject to SPOT and RT-qPCR detection in parallel. miR-155 was used as biomarker for breast cancer^73^, while miR-21 was used as biomarker for gastric and prostate cancer, which were reported to be upregulated in patients^74, 75^. Noted that expression levels of many miRNAs are highly dynamic and heterogenous, thus a measurement and direct comparison of miRNA abundance may not accurately reflect the pathological state of the samples, therefore, measuring a housekeeping gene at the same time is essential for miRNA-based diagnosis assays^76^. Herein, miR-16 was selected as the housekeeping gene to serve as an internal reference^77^. We collected serum samples from 10 breast cancer patients, 12 gastric cancer patients, and 10 prostate cancer patients along with a number of samples from healthy donors (Table S2). The relative expression level of miR-155 and miR-21 against the housekeeping gene miR-16 as detected by SPOT were depicted in Fig. 5a. As expected, patients of breast cancer (B.C.), gastric cancer (G.C.), and prostate cancer (P.C.) exhibited a relatively higher expression of miR-155 or miR-21 comparing to healthy donors (H.D.). The diagnostic performance of these miRNAs as detected by SPOT was quantitatively evaluated by receiver operating characteristic (ROC) curve analysis (Fig. 5b). SPOT detection of miRNAs yielded excellent area under the curve (AUC) values for cancer diagnosis, with 0.854 for B.C., 0.775 for G.C., and 0.880 for P.C., whose diagnosis power are slightly lower but comparable to the results of RT-qPCR when analyzing the miRNA samples from the same group of patients (Fig. S18, Fig. 5c).

**Fig. 5.**
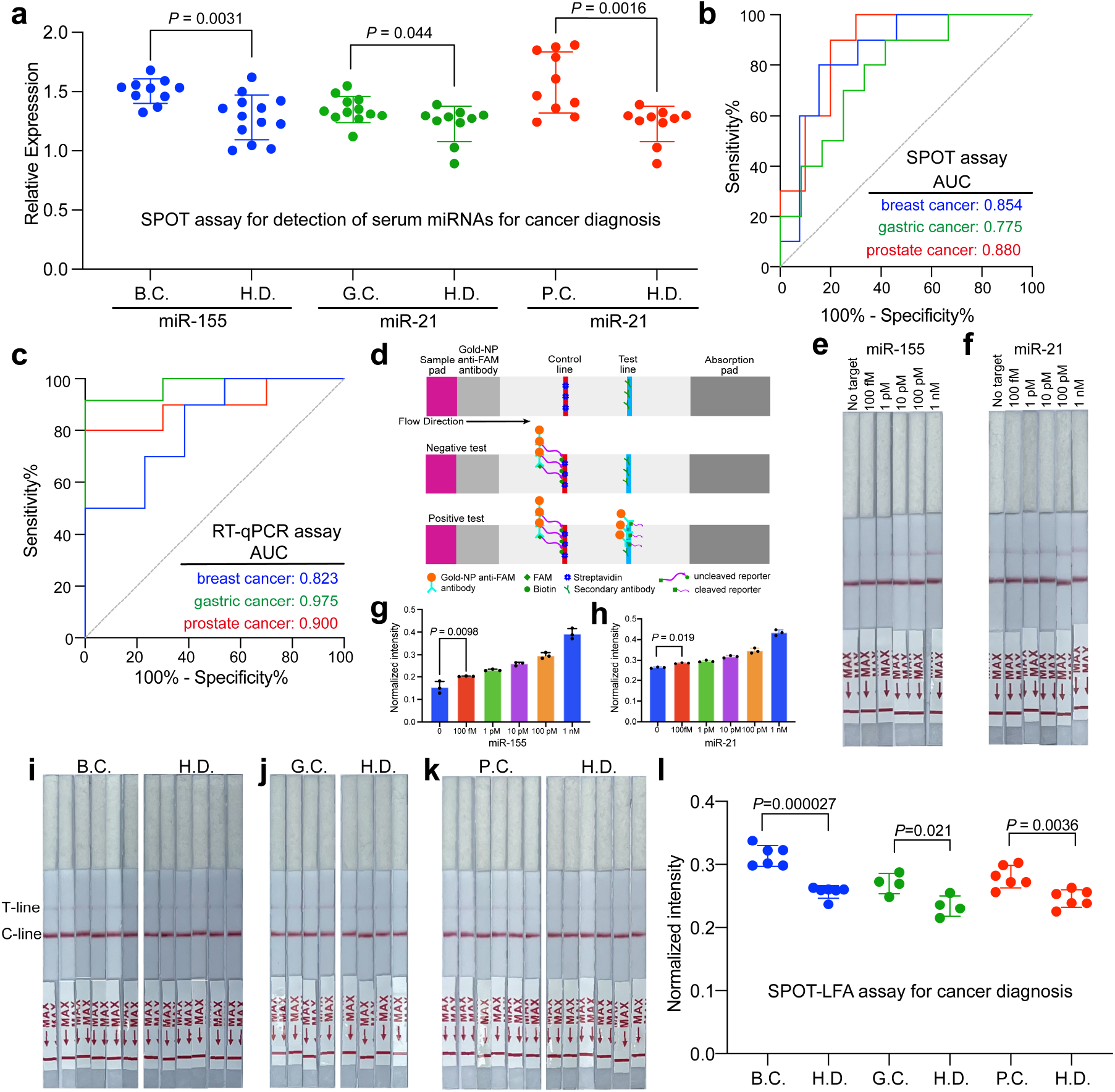
Detection of serum miRNAs for cancer diagnosis. **a**, Scatter dot plots showing the relative expression level of miR-155 or miR-21 against miR-16 for breast cancer patients (n=10), gastric cancer patients (n=12), and prostate cancer patients (n=10), along with a number of healthy donors. The middle line and error bar represent the mean and SD, respectively. **b**, ROC curve and AUC analysis for breast cancer, gastric cancer, and prostate cancer diagnosis by SPOT. **c**, ROC curve and AUC analysis for multiple cancer diagnosis by using RT-qPCR. **d**, Principle of lateral flow assay for visual detection of miRNA targets by using SPOT. **e**,**f**, Representative photos for SPOT-LFA detection of synthetic miR-155 (**e**) and miR-21 (**f**) at variable concentrations. **g**,**h**, Bar graphs for the intensity of test lines for miR-155 (**g**) and miR-16 (**h**). Error bar, 1 s.d. (n = 3). **i-k**, SPOT-LFA detection of serum miRNAs for molecular diagnosis of breast cancer (**i**, n=6), gastric cancer (**j**, n=4), or prostate cancer (**k**, n=6). **l**. Scatter dot plots for discriminating cancer patients from healthy donors by SPOT-LFA assay. The middle line and error bar represent the mean and SD, respectively. *P* values were calculated by two-tailed Student’s t-test.

The simplicity and robustness of SPOT assay enables it adaptable for POCT applications. Herein, for demonstration, we coupled SPOT with LFA for the development of SPOT-LFA assay towards instrument-free molecular diagnosis of cancers. Standard commercially available LFA test strips can be readily adapted for SPOT-LFA (Fig. 5d). Visual detection of synthetic miRNAs of various concentrations by SPOT-LFA were presented in Fig. 5e and Fig. 5f, whose test line intensity were quantified and then normalized based on intensity of control lines (Fig. 5g,5h), showing good linearity between miRNA concentrations and measured intensity. The calculated LOD are 968 fM and 32 fM for miR-155 and miR-21, respectively (Fig. S19), lower than above fluorescent SPOT assays most likely due to reduced sensitivity in LFA visualization. We then applied SPOT-LFA to patient samples that were clinically diagnosed with cancers or healthy controls (Fig. 5i-k), which revealed good diagnosis performance by successfully discriminating cancer patients from healthy donors (Fig. 5l). Overall, these experimental assessments with clinical patient samples validated the high sensitivity, specificity, and ease of operation of our SPOT-based assay for miRNA detection towards precise cancer diagnosis.

### Preamplification-free detection of SARS-CoV-2 RNA

We next sought to expand the application capability of SPOT to other biological nucleic acids such as SARS-CoV-2 RNAs. Unlike miRNAs, SARS-CoV-2 RNA has a much lower abundance (in the attomolar range) in clinical samples^11^, which thus demands a prerequisite preamplification for most detection assays of insufficient sensitivity. As demonstrated above, the best LOD of SPOT for miRNA detection is in the range of femtomolar, which still falls short for detecting clinical SARS-CoV-2 RNA. To boost the sensitivity of SPOT, inspired from a recent work on Cas13a-based SARS-CoV-2 RNA detection,^78^ we developed a multiplex SPOT assay for combinatory detection of various distinct gene sites on the same genome of SARS-CoV-2 (Fig. 6a). We designed a number of fourteen SPOTs targeting the Orf1ab gene and N gene sites on the SARS-CoV-2 RNA genome (Fig. 6b, Fig. S20). Prior to multiplex combination of SPOTs, we first evaluated and screened the performance of individual SPOT designs by detecting a short synthetic nucleic acid with the same sequence of its corresponding gene site on the SARS-CoV-2 genome (Fig. S21). The top 5 best SPOT designs were then selected for multiplexing study, showing that triplex SPOT assays exhibit drastically enhanced detection capability than that of duplex and single SPOT assays (Fig. 6c, S22). After selecting the best triplex SPOT, we went on to examine its detection sensitivity against corresponding synthetic RNA targets, which showed to have a calculated LOD of 1.90 aM (Fig. 6d), largely improved from the previous single SPOT assays for miRNAs. Furthermore, triplex SPOT assay was demonstrated capable of detecting in vitro transcribed (IVT) SARS-CoV-2 reference RNA with concentration as low as 130 aM (Fig. 6e). These experiments suggested that triplex SPOT assay shall be able to realize sensitive detection of SARS-CoV-2 RNA in clinical samples without involving preamplifications.

**Fig. 6.**
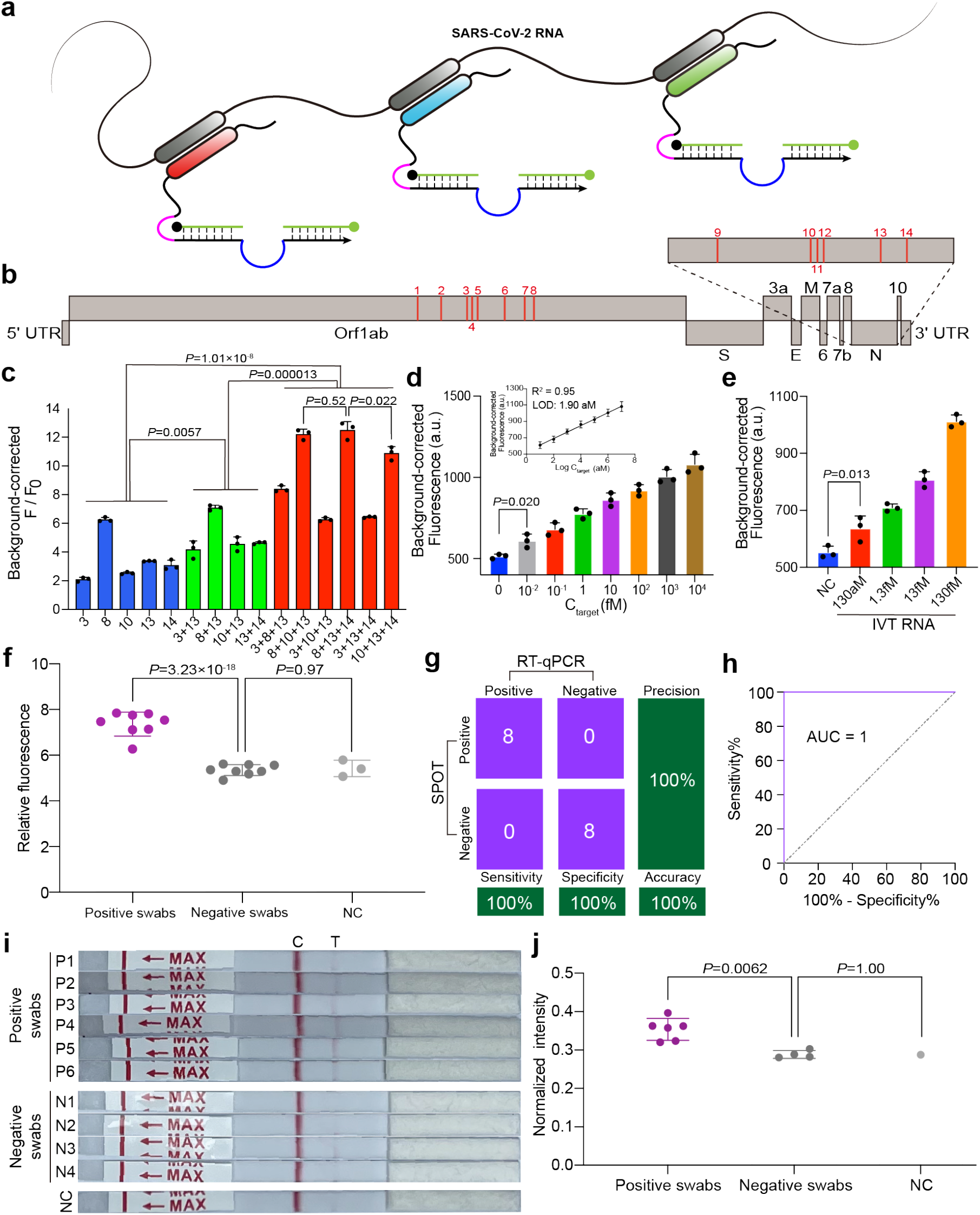
Performance of SPOT for preamplification-free detection of SARS-CoV-2 RNA. **a**, Schematic of detecting SARS-Cov-2 RNA with combinatory use of multiple SPOT systems simultaneously. **b**, A number of 14 sites within the orf1ab and N genes are selected and screened for multiplex SPOT. **c**, Combinatory use of SPOTs enable enhanced detection capability. Blue: single SPOT. Green: duplex SPOT. Red: triplex SPOT. SPOT: total 12 nM, target: 500 pM for each target, reporter: 500 nM. **d**, Examining the detection sensitivity on synthetic SARS-Cov-2 RNA targets by using triple-SPOT (10, 13, and 14) systems in one-pot detection without preamplifications. Inset: Corresponding linear curve between fluorescence and logarithm of target concentrations. A LOD of 1.90 aM is calculated from this curve. SPOT: 4 nM for each SPOT, reporter: 500 nM. **e**, Detection of IVT SARS-Cov-2 RNA by triplex SPOT. SPOT: 4 nM for each SPOT, reporter: 500 nM. **f**, Detection of SARS-CoV-2 RNA that pre-extracted from sixteen nasopharyngeal swabs by using SPOT assay. **g**, Confusion matrix analysis of the sixteen clinical samples as detected by SPOT and RT-qPCR. **h**, ROC curve of SPOT for diagnosing SARS-CoV-2 infection. The clinical samples were correctly classified with AUC = 1 (95% CI: 1.0000 to 1.0000). **i**, SPOT-LFA assay for detection of SARS-CoV-2 RNA from clinical samples. **j**, Scatter plot of SPOT-LFA assay for discriminating positive swabs from negative swabs. All measurements are mean ± SD with n = 3. *P* values were calculated by two-tailed Student’s t-test.

We subsequently used the triplex SPOT assay to detect SARS-CoV-2 RNA from clinical nasopharyngeal swabs of patients with known infection status as confirmed by RT-qPCR (Fig. 6f, Table S2). Out of 16 patients, SPOT was able to correctly identify all 8 positive samples and 8 negative samples, exhibiting a calculated sensitivity of 100%, specificity of 100% (Fig. 6g). The accuracy of this assay was 100% with an AUC of 1.00 (95% CI: 1.0000 to 1.0000, Fig. 6h). Furthermore, SPOT-LFA assay can also be readily applied to preamplification-free detection and visualization of clinical SARS-CoV-2 RNA samples from patients (Fig. 6i), which can discriminate positive from negative samples and negative controls (Fig. 6j). In brief, we have realized preamplification-free, one-pot, one-step, and accurate detection of SARS-CoV-2 infections from clinical samples by using multiplex SPOT assay.

## Discussion

Circulating nucleic acids in biofluids are emerging biomarkers for a variety of diseases, whose abundance, however, are extremely low, posting great hurdles for direct detections. Amplification of targeting nucleic acids generally accompany with detection assays by using PCR or other isothermal methods (e.g., LAMP, RPA, RCA) to compensate its insufficiency in sensitivity. Nevertheless, amplification not only increases the chances of false positive detection or leakage of amplicons, but also necessitates the need of centralized laboratory, sophisticated instruments^8, 79^, trained personnel, and prolong the overall protocol due to its serial nature. Efforts in developing amplification-free assays typically focus on using specialized devices such as electrochemical sensors to enhance sensitivity^80^, which are relatively hard to be fabricated. Others focus on engineering CRISPR-Cas systems to boost its detection sensitivity, such as by using Cas systems against various sites within a long nucleic acid target^78^, or by using cascade designs of multiple Cas endonucleases^81^. Though these CRISPR-Cas-based amplification-free methods have achieved great success in detecting long viral RNA targets, which may fall shorthanded on detecting short miRNAs due to crRNA sequence design constrains. In addition, the use of expensive Cas endonuclease of short shelf life renders this assay less cost-effective.

Here in this report, our SPOT assay enables highly sensitive detection of single-stranded nucleic acids with no need of amplifications. SPOT is an allosteric DNAzyme-based biosensing molecular system, which is designed and constructed from a single DNA strand. We conducted systematic examination on design parameters (e.g., locking domain length, detection loop length, substrate constitution *etc*.) and experimental conditions (e.g., temperature, ionic strength, reaction time *etc*.) for achieving optimal detection performance of SPOT. Furthermore, the mechanistic activation pathway of SPOT upon target recognition was experimentally studied and computational validated by NUPACK and oxDNA MD simulations. Such a comprehensive understanding on the effects of design parameters and molecular mechanisms may shed light on designing DNAzyme-based biosensing systems in the future towards a large diversity of molecular targets including nucleic acids, proteins, small molecules, and beyond. The LOD of SPOT assay can reach to as low as tens of femtomolar for miRNA, and to single-digit attomolar for SARS-CoV-2 RNA when using multiplex SPOTs. In addition to its high sensitivity, SPOT exhibits potent specificity on discriminating nucleic acid targets of single nucleotide mutations. Due to sequence-specific cleavage of substrates by the DNAzyme, SPOT shows excellent orthogonality in target detection and signal readout, enabling multiplexing capability for sensing various targets in a one-pot system at the same time. SPOT is highly cost-effective given it is fully constructed from synthetic DNA molecules, which spares the need of protein-based costly enzymes. The estimated material cost for one SPOT assay can be as low as ∼0.02 dollars at a research scale, which is significantly lower than reported CRISPR-Cas-based assays (Table S3)^31^. Lastly, SPOT can be readily adapted for POCT-based diagnostics such as LFA-based modalities for detection of miRNAs towards cancer diagnosis, or for detection of SARS-CoV-2 RNA towards viral infection diagnosis. Such a combination of sensitive and specific analytical performance, one-pot and one-step operational protocol, low cost, and POCT capability enables SPOT a substantially improved assay comparing to existing DNAzyme or CRISPR-based assays for nucleic acid targets in clinical settings.

## Data Availability

All data produced in the present study are available upon reasonable request to the authors

## Acknowledgements

The authors thank the supports from the National Key Research and Development Program of China (2021YFA0910100), from the National Natural Science Foundation of China (21974086), and from the Innovative Research Team of High-Level Local Universities in Shanghai (SHSMU-ZLCX20212602).

