## Supplementary information for "Allosteric DNAzyme for sensitive detection of nucleic acids for molecular diagnosis"

#### Materials and Methods.

#### Overview of Assays.

Sequences of all oligonucleotides were shown in Table S1. Oligonucleotide strands were purchased from Sangon Biotech or TaKaRa or Bioligo, and all strands were dissolved in depc (diethylpyrocarbonate) treated water (Sangon, B501005) and quantitated by a Thermo Scientific Nanodrop Spectrophotometer with final concentration of 20 μM.

All assays for 10-23 SPOT and free-form 10-23 DNAzyme were conducted in 1×10-23 buffer: 4 × TE buffer (40 mM Tris, 4 mM EDTA, pH = 7.5) with 150 mM $Mg^{2+}$ except assays for optimization. Moreover, all assays for 13PD1 SPOT and free-form 13PD1 DNAzyme were conducted in 1×13PD1 buffer: HEPES buffer (70 mM HEPES, pH = 7.5) with 1 mM $Zn^{2+}$, 20 mM $Mn^{2+}$, 40 mM $Mg^{2+}$ and 150 mM $Na^{+}$.

For all SPOT assay, firstly, all self-locked DNAzymes were denatured at 95 °C for 3 min and cooled down in their corresponding buffer under a ramp annealing protocol (from 95 °C to 4 °C) over a period of 35 min. Then these DNAzymes were incubated with targets and reporters at 28 °C for the 10-23 SPOT and at 37 °C for the 13PD1 SPOT with fluorescence measurements. For fluorescence measured by a multifunctional microporous plate detector (Biotek Synergy H1), all reactions were conducted on black 96-well plates (WHB-96-02). For fluorescence measured by fluorescent real-time quantitative thermal cycling apparatus (cobas, z480), all reactions were conducted on PCR 8-Tube (Sangon, F602004). To obtain the background-corrected fluorescence values, the fluorescence values obtained from reactions carried out with only the reporter and buffer were subtracted.

#### Cleavage of substrates by free-form DNAzyme

For 10-23 DNAzyme: 500 nM R-L, D/R-L, D/R-MB3 and D/R-MB4 were incubated with or without 10 nM free-form 10-23 DNAzyme, respectively, at 28 ℃ in total volume 100 μL on a black 96-well plate. Fluorescence measurements were taken every 70 seconds using an excitation wavelength of 485 nm and an emission wavelength of 528 nm with a gain of 100. The measurements were recorded for a duration of 2 hours.

For 13PD1 DNAzyme: 1 μM 13PD1 reporter were incubated with or without 100 nM, 10 nM, 1 nM, 100 pM and 10 pM free-form 13PD1 DNAzyme, respectively, at 37 ℃ in total volume 100 μL on a black 96-well plate. Herein, since 13PD1 DNAzyme is significantly different from 10-23 DNAzyme in terms of catalytic core, reporter and catalytic cofactor, cleavage temperature for 13PD1 was the same as previous reports[1,2]. Fluorescence measurements were taken every 5 minutes using an excitation wavelength of 485 nm and an emission wavelength of 528 nm with a gain of 100. The measurements were recorded for a duration of 1 hour.

**Screening and examining detection capability of SPOT designs for synthetic miRNA detection**

For screening SPOT design, 100 nM DNAzyme was first annealed in the 1×10-23 buffer. Then 10 μL annealed DNAzyme (100 nM, 1×10-23 buffer), 10 μL miR-155 (100 nM, 1×10-23 buffer) and 10 μL reporter (5 μM, 1×10-23 buffer) were added to 70 μL 1×10-23 buffer to reach 100 μL. Fluorescence was measured for every 70 s in a multifunctional microporous plate detector during 28 °C incubation for 2 h (excitation wavelength: 485 nM; emission wavelength: 528 nM; Gain: 100).

For examining the detection capability, after annealing, the 10 μL DNAzyme (100 nM, 1×10-23 buffer) was mixed with 10 μL targets (10×final concentration, 1×10-23 buffer) and 10 μL reporter (5 μM, 1×10-23 buffer) in 70 μL 1×10-23 buffer for 2 h incubation at 28 °C. Fluorescence was measured after the incubation.

**Optimizing experimental conditions for SPOT constructed from 10-23 DNAzyme**

To achieve fast and sensitive detection of miRNA and SARS-CoV-2 RNA, we took several optimizations for 10-23 SPOT based on 500 pM miR-155. Since concentration of ion is highly related to cleavage activity of DNAzyme[3], concentration of $Mg^{2+}$ in the 10-23 buffer was first optimized. By increasing final concentration of $Mg^{2+}$ from 10 mM to 150 mM with maintaining 10 nM DNAzyme, 500 nM reporter and 2 h incubation at

28 °C fluorescence was measured at 2 h for with (F) and without ($F_0$) the target. Moreover, since the incubation temperature is highly related to SNR, incubation temperature (25 °C, 28 °C, 31 °C, 34 °C, 37 °C) was optimized with maintaining 10 nM DNAzyme, 500 nM reporter, 150 mM $Mg^{2+}$ in the 10-23 buffer and the 2 h incubation. Furthermore, final concentration of DNAzyme was optimized with maintaining 150 mM $Mg^{2+}$ in the 10-23 buffer and the 2 h incubation at 28 °C. Finally, to fulfill fast detection, its duration was optimized by monitoring fluorescence with and without the target for 5 h 30 min in the 10-23 buffer with the best optimized conditions. We chose 2 h as the detection duration. Fluorescence was measured for every 70 s (excitation wavelength :485 nM; emission wavelength :528 nM; Gain: 100). All measurements were taken on black 96-well plates with 100 μL volume.

**SPOT constructed from 13PD1 DNAzyme for miRNA detection**

1 μM DNAzyme was first annealed in the 1×13PD1 buffer. Then 10 μL DNAzyme (1 μM, 1×13PD1 buffer), 10 μL DNA mimic of miR-155 (1 μM, 1×13PD1 buffer) and 10 μL reporter (10 μM, 1×13PD1 buffer) were added to 70 μL 1×13PD1 buffer to reach 100 μL. Fluorescence was measured in a multifunctional microporous plate detector after 2 h incubation at 37 °C (excitation wavelength :485 nM; emission wavelength :528 nM; Gain: 100).

**NUPACK simulations**

All simulations were conducted in NUPACK with DNA nucleic acid type at 28 °C[4]. For DNAzyme simulation, number of strand species and maximum complex size were both 1. For DNAzyme / target interaction simulation, number of strand species was 2, and maximum complex size was 3.

**MD simulations of structures of the SPOT**

To computationally predict structures of SPOT and take SPOT mechanism studies, we performed molecular dynamics (MD) simulations using the oxDNA coarse-grained model of nucleic acids[5,6]. The simulation objects include all SPOT designs with different length of locking domain (Fig. S3) and after addition of miR-155 DNA mimic target for SPOT-7+5. oxDNA model has been employed for simulating structures by comprehensively considering mechanical and thermodynamic characteristics of single- and double-stranded nucleic acids, exhibiting favorable consistency with experimental observations. Moreover, this method enables simulations with a sufficiently long timescale by using computationally accessible resources[7,8].

All simulated objects were designed in oxView[5]. All simulations were conducted in the oxDNA program using default procedures at 28 °C by DNA interaction type. Distance distribution between any two bases were calculated by oxDNA program and graphs were plotted by Graphpad Prism 9.

**Native polyacrylamide gel electrophoresis (PAGE)**

All samples for gel electrophoresis were prepared in the same way as the fluorescent assays. 200 nM annealed SPOT was incubated with 200 nM miR-155 for and 5 μM reporter at 28 °C for 2 h. 10 μL for each lane were subjected to 15%-page gel electrophoresis at 120 V for 2.5 h in an ice−water bath. Gels were prepared with 0.5 × TBE buffer containing 10 mM $MgCl_2$ and left to solidify at room temperature for at least 1 h before using. GelRed (41003, Biotium) was used for postelectrophoresis page gel staining. The gel was scanned using an Amersham Imager 680 RGB (GE Healthcare).

**Serum miRNA extraction and detection by SPOT for cancer diagnosis**

All serum samples were obtained from Renji Hospital, Shanghai Jiao Tong University School of Medicine (Shanghai, China). The study was approved by the Ethics Committee at Renji Hospital, School of Medicine, Shanghai Jiao Tong University (KY2023-003-B). All methods were performed in accordance with these approved guidelines. Total RNA in serum samples was extracted using the miRNeasy Serum/Plasma Kit from Qiagen of Canada. All samples were purified according to the manufacturer’s instructions. The eluted RNAs were stored in nuclease-free water at −80 °C until needed.

100 nM DNAzyme was first annealed in the 1×TE, 150 mM $Mg^{2+}$ buffer. Then 2 μL DNAzyme (100 nM, 1×TE, 150 mM $Mg^{2+}$), 2 μL extracted serum miRNAs and 2 μL (5 μM, 1×TE, 150 mM $Mg^{2+}$) reporter were added to 14 μL 1×10-23 buffer to reach 20 μL. Fluorescence was measured by fluorescent real-time quantitative thermal cycling apparatus through FAM channel after 2 h incubation at 28 °C.

**Analysis of relative expression of miRNAs from serum samples with RT-qPCR.**

The cDNA was prepared by miRcute Plus miRNA First-Strand cDNA Synthesis Kit (TIANGEN, KR211). In this step, 20 μL of the RT system included 10 μL of 2× miRNA RT reaction buffer, 2 μL of miRNA RT Enzyme Mix, 2 μL of eluted RNA and 6 μL of depc treated water. The RT procedure was 42 °C for 60 min and 95 °C for 3 min. The cDNA solution was diluted by 3× before being added to qPCR system. qPCR analysis of miR-155, miR-21 and miR-16 was performed with miRcute Plus miRNA qPCR Kit (SYBR Green) (TIANGEN, FP411). 20 μL of the qPCR system included 10 μL of 2× miRcute Plus miRNA Premix (STBR&ROX), 0.4 μL 10 μM of forward primer (TIANGEN, CD201/CD202), 0.4 μL 10 μM of reverse primer, 2 μL of the cDNA template and 7.2 μL of depc treated water. The qPCR procedure was 95 °C for 15 min, 45 cycles of 94 °C for 20 s and 60 °C for 34 s. In brief, the relative expression of miR-155 and miR-21 from serum samples was evaluated by referring to the expression of miR-16 using the $2^{-\Delta\Delta Ct}$ method.

**Lateral flow assay**

The LFA strips were purchased from Shanghai Toolman Biotech. Black hole quencher 1 (BHQ1) modified on reporters were replaced by biotin (5' FAM-CACTTCTrArUTTCCCC-Biotin 3'). Concentration of all reagents and procedures were the same as the fluorescent assays except 4 h incubation at 28 °C. 50 μL SPOT reactions were mixed with 50 μL lateral flow test buffer. After mixing, the strips were immersed into the solution with 5 min incubation. Then strips were taken out. Photos were obtained by iPhone 12 camara with default parameters. The LFA images were analyzed by imageJ software with following procedure: transform images into 8-bit type;

subtract background; set measurements to take analysis of integrated density; invert color; use the rectangle tool to obtain the integrated density of the test line. Integrated density was normalized with averaged control line intensity.

**Screening and examining detection capability of SPOT designs for synthetic SARS-CoV-2 RNA detection**

All assays for the screening the sensor was assembled by final concentrations of 10 nM DNAzyme and 500 nM reporter with 10 nM target in 1×10-23 buffer. Procedure was the same as screening assays for miRNA detection. For the combinatory use of SPOTs all DNAzymes and targets were incubated together with one-$X^{th}$ (in X DNAzymes combinations) concentration for each DNAzyme to keep the 12 nM total concentration and 500 pM for each target. For kinetic assays, fluorescence was measured every 70 s (excitation wavelength: 485 nM; emission wavelength: 528 nM; Gain: 100). For synthetic SARS-CoV-2 RNA detection, the process is the same as synthetic miRNA detection.

**SARS-CoV-2 RNA extraction and detection by SPOT for SARS-CoV-2 diagnosis**

All SARS-CoV-2 clinical samples were obtained from Renji Hospital, Shanghai Jiao Tong University School of Medicine (Shanghai, China). The study was approved by the Ethics Committee at Renji Hospital, School of Medicine, Shanghai Jiao Tong University (KY2022-192). The nucleic acid was isolated from a 200 μL swab sample using an automated nucleic acid extraction instrument (Smart Lab Assist) and the accompanying reagents (Taiwan Advanced Nanotech, Taiwan, China) following the manufacturer's instructions. The extracted nucleic acid was then eluted in a 50 μL depc treated water.

120 nM DNAzyme-10, DNAzyme-13 and DNAzyme-14 was first annealed in the 1×TE, 150 mM $Mg^{2+}$ buffer, respectively. Then the three DNAzyme was mixed based on 1:1:1 volume ratio. 2 μL the mixed DNAzyme (100 nM, 1×TE, 150 mM $Mg^{2+}$), 2 μL extracted serum miRNAs and 2 μL reporter (5 μM, 1×TE, 150 mM $Mg^{2+}$) were added to 14 μL 1×10-23 buffer reach 20 μL. Fluorescence was measured by fluorescent real-

time quantitative thermal cycling apparatus through FAM channel after 2 h incubation at 28 °C.

**Statistical analysis**

Equation of linear regression for background-corrected fluorescence and logarithm of target concentrations was conducted by Graphpad Prism. For comparing the patient and control groups, a two-tailed unpaired t-test was conducted. ROC analyses were used to draw ROC curve and determine AUC values by Graphpad Prism. Distribution analyses by Graphpad Prism were used to draw distance distribution between two nucleotides from oxDNA simulation.

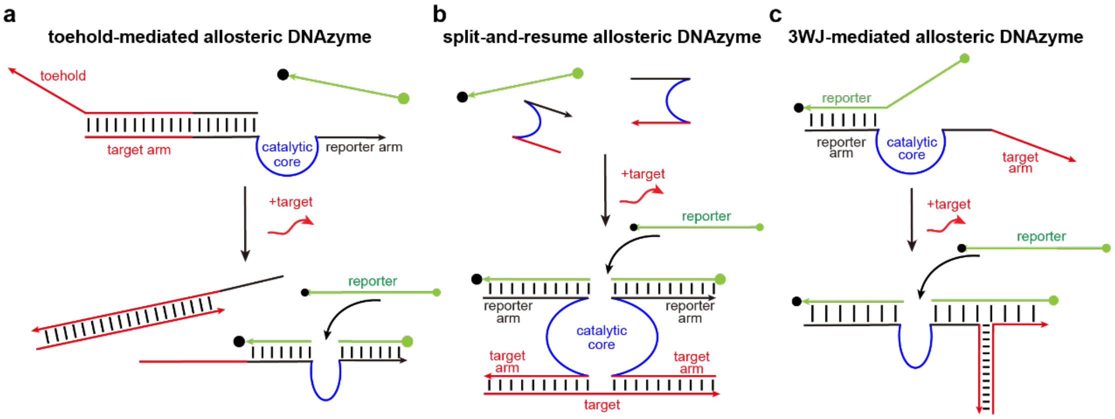

**Fig. S1 Conventional allosteric DNAzyme biosensors with multicomponent molecular complexes. a,** Toehold-mediated allosteric DNAzyme regulation, achieved by inhibiting or activating the DNAzyme through an inhibitor strand that binds to a toehold region[9]. **b,** split-and-resume allosteric DNAzyme by splitting and resuming the catalytic core upon target binding[10–12]. **c,** 3WJ-mediated allosteric DNAzyme by target-induced stabilization of DNAzyme-substrate complex[13].

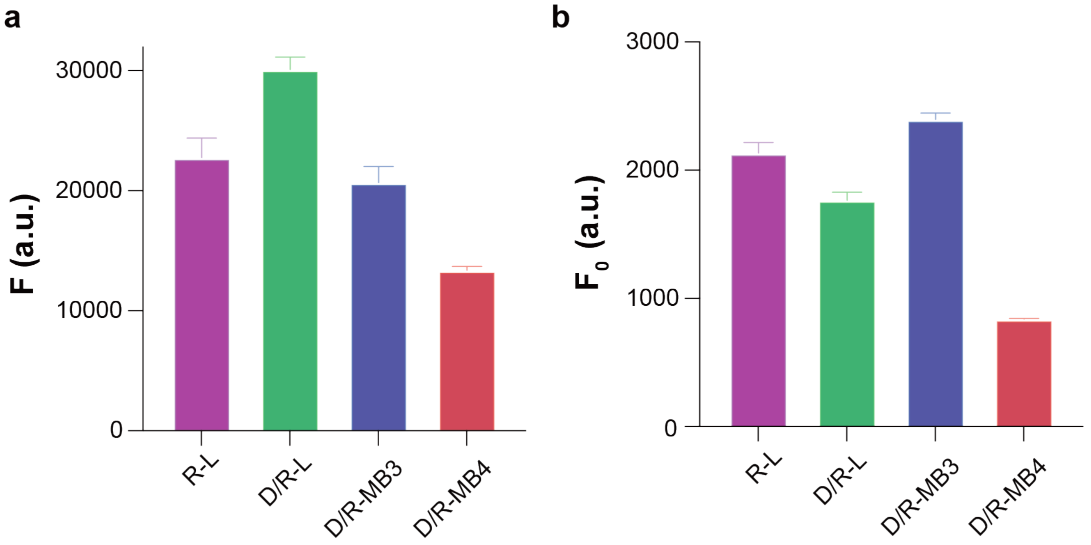

**Fig. S2 Reporter optimization.** Fluorescence of **a,** signal (F, with DNAzyme) and **b,** background noise (F0, without DNAzyme) after 2 hours of cleavage against various reporters by free-form 10-23 DNAzyme. All experimental measurements are mean ± standard deviation (SD) with n = 3.

### Design and Screening of SPOT

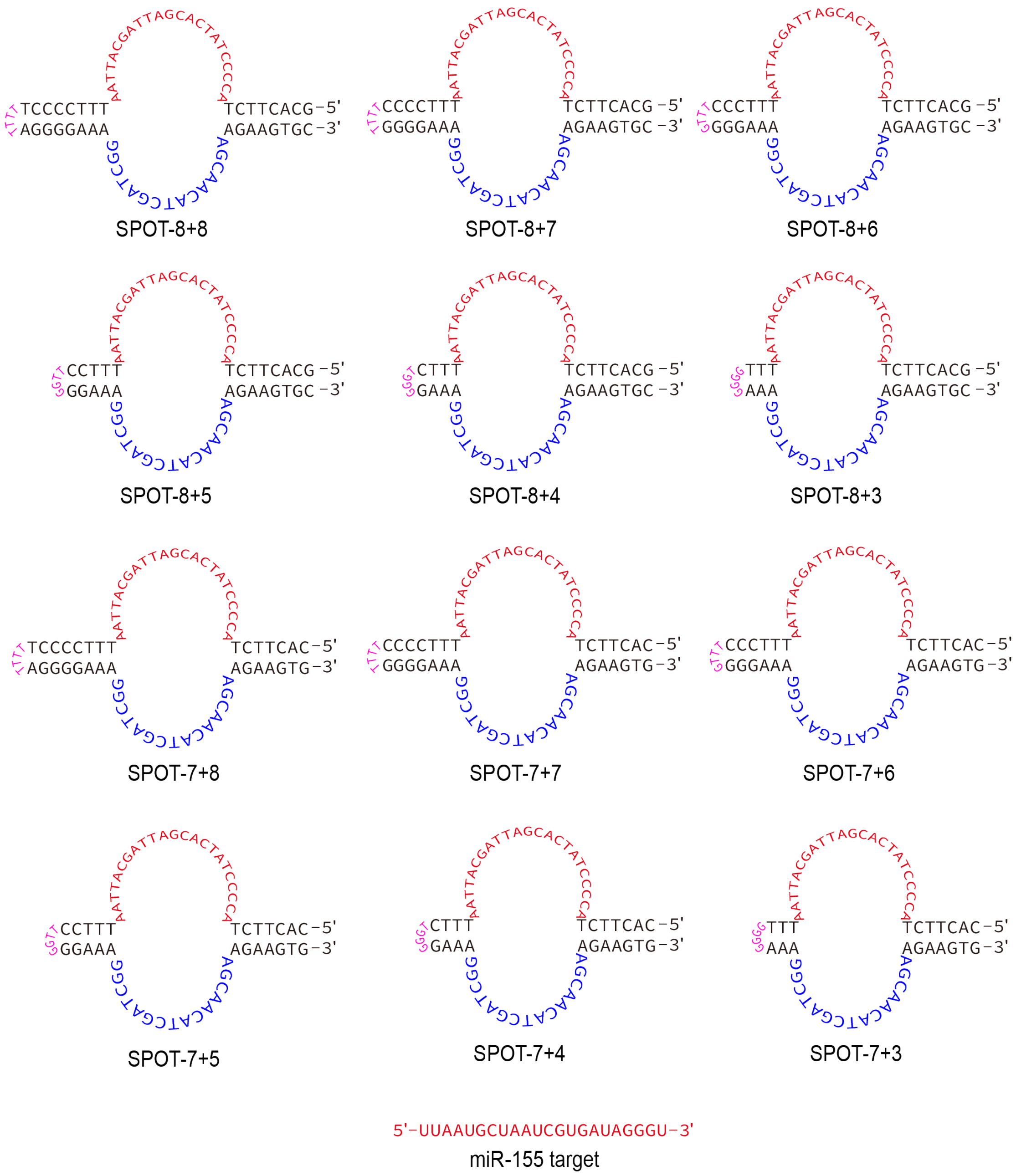

**Fig. S3 Design of SPOT constructed from 10-23 DNAzyme with domain1 (7-bp or 8-bp) and domain2 (3-bp to 8-bp).**

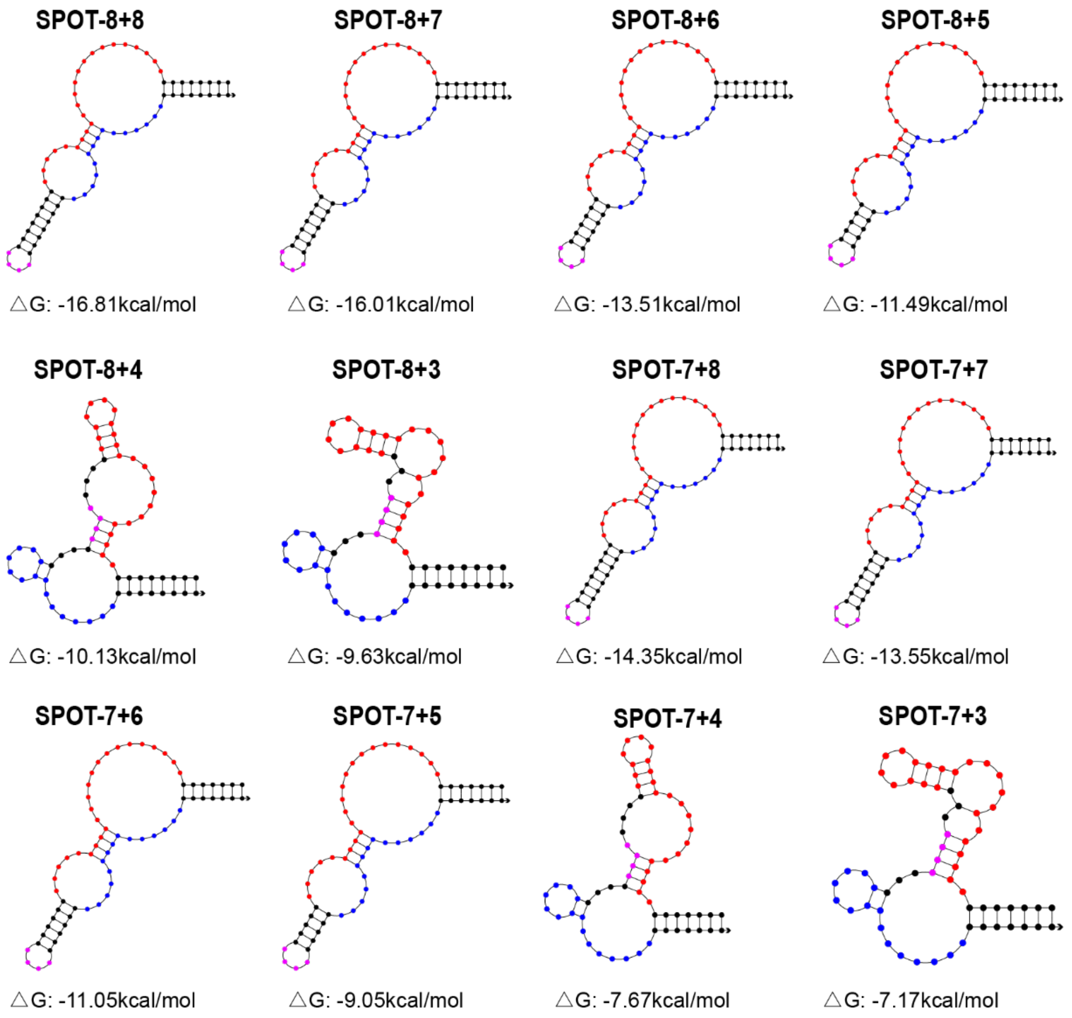

**Fig. S4 Simulated secondary structures and Gibbs free energy change of SPOTs by using NUPACK[4].**

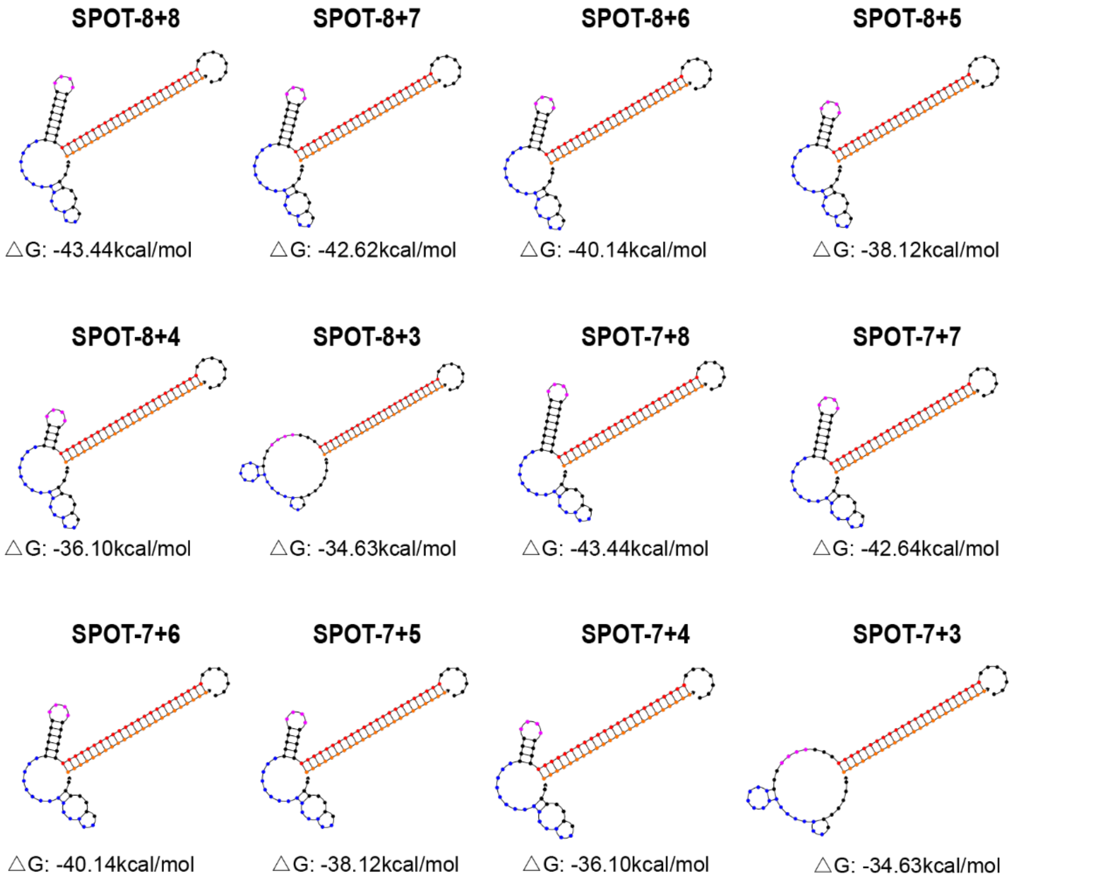

**Fig. S5 Simulated secondary structures and Gibbs free energy change of SPOT-target complexes by using NUPACK[4].**

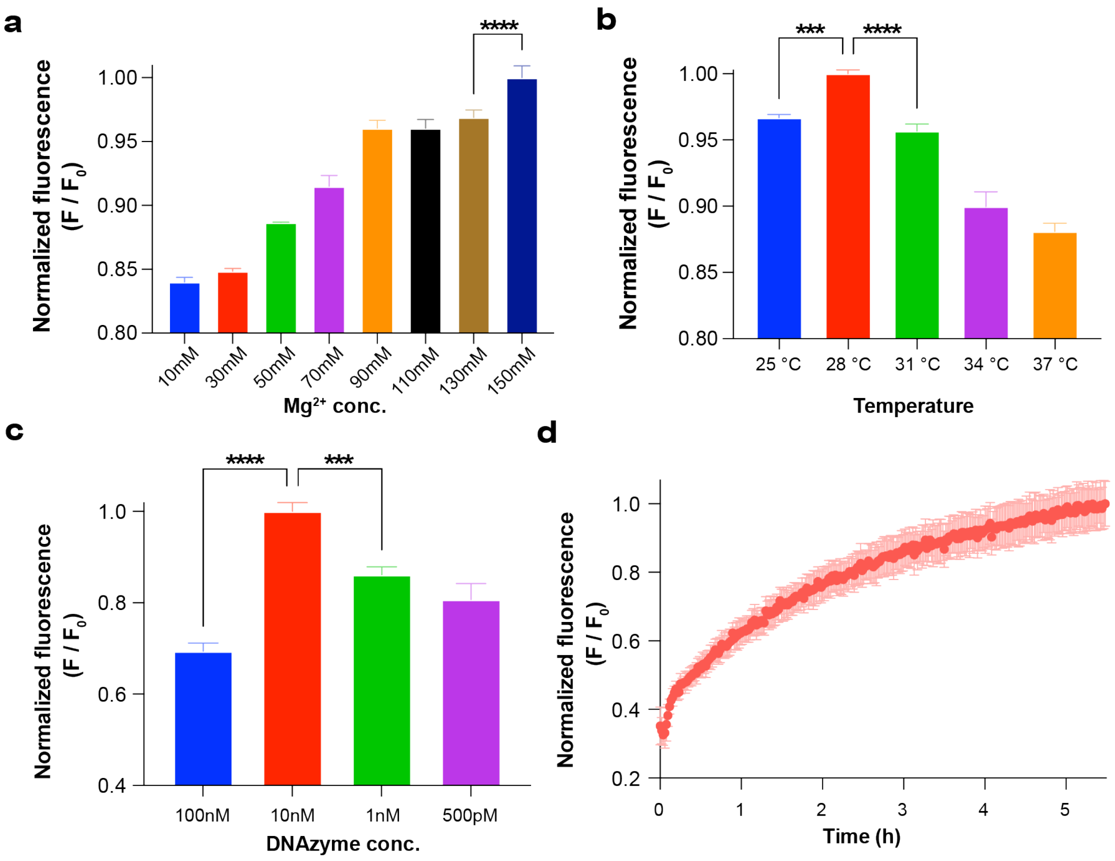

**Fig. S6 Optimization for SPOT detection. a,** Optimization of concentration of $Mg^{2+}$ in the reaction buffer. **b,** Optimization of cleavage temperature. **c,** Optimization of concentration of DNAzyme. **d,** Optimization of reaction duration. DNAzyme, 10nM; target. 500 pM; reporter, 500 nM. All experimental measurements are mean ± standard deviation (SD) with n = 3. $^{***}P < 0.001$ and $^{****}P < 0.0001$ by T-test.

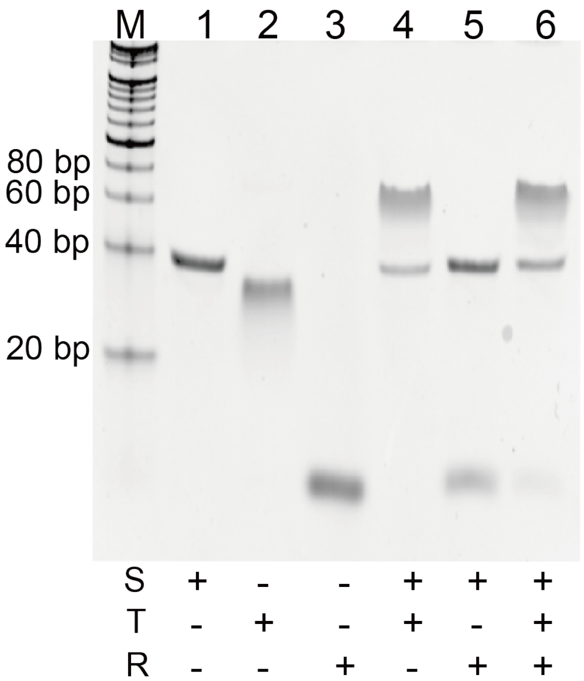

**Fig. S7 Native PAGE analysis of SPOT.** Lane M: 20 bp DNA ladder; Lane 1: SPOT (S); Lane 2: RNA target (T); Lane 3: RNA reporter (R); Lane 4: mixture of SPOT and target RNA at 1: 1 molar ratio; Lane 5: mixture of SPOT and RNA reporter at 1: 25 molar ratio; Lane 6: mixture of SPOT, target RNA, and RNA reporter at 1: 1: 25 molar ratio. All samples were incubated for 2 hrs before gel analysis.

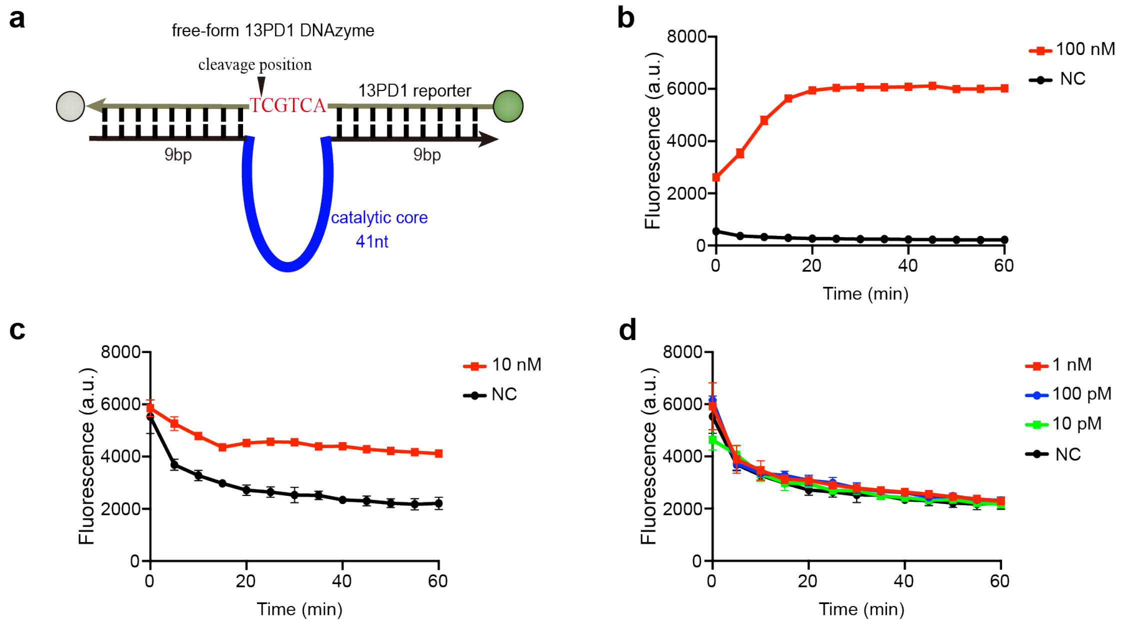

**Fig. S8 The properties of 13PD1 DNAzyme. a,** Schematic of hydrolysis of reporter by free-form 13PD1 DNAzyme. Red character: unpaired nucleotides. Green ball: FAM fluorophore. Gray ball: Black Hole Quencher 1. Bold blue loop: catalytic core of 13PD1 DNAzyme. **b, c, d** Kinetics of 13PD1 cleavage. 100 nM (Red line, b), 10nM (Red line, c), 1 nM (Red line, d), 100 pM (Blue line, d) and 10 pM (Green line, d) unlocked-13PD1 DNAzyme plus 1 μM 13PD1-reporter (FQ), respectively. Black line: 1 μM 13PD1-reporter (FQ). Fluorescence was recorded every 5 mins. All experimental measurements are mean ± standard deviation (SD) with n = 3.

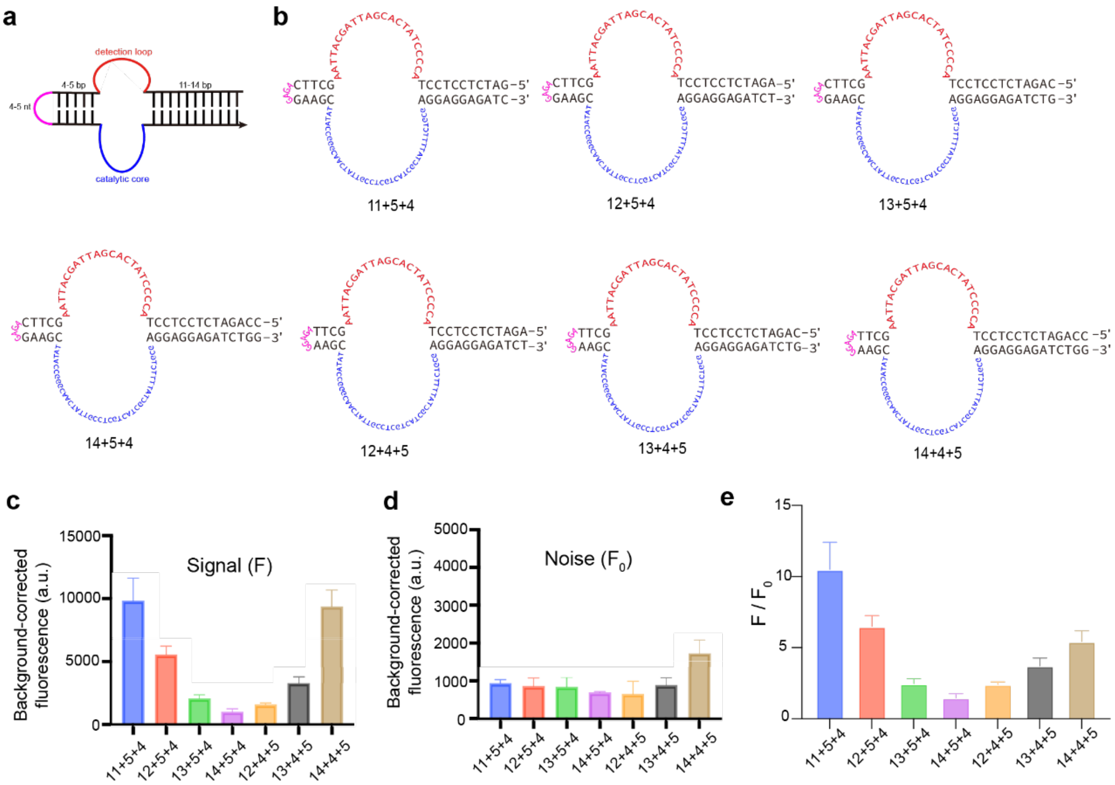

**Fig. S9 13PD1 SPOT. a,**Schematic designs of SPOT based on 13PD1 DNAzyme. **b,** Design of SPOT constructed from 13PD1 DNAzyme with domain1 (11-bp to 14-bp), domain2 (4-bp or 5-bp) and loop (4-nt or 5-nt). **c, d,** c, signal (F, with target) and d, background noise ($F_0$, without target) after 1 hour of cleavage against 13PD1 reporters by all 13PD1 SPOTs. **E,** Fluorescence signal-to-noise ratio (SNR) of SPOTs for the detection of DNA-mimic miR-155 after 60 mins of incubation. SPOT-11+5+4 design exhibits the highest SNR. All experimental measurements are mean ± standard deviation (SD) with n = 3.

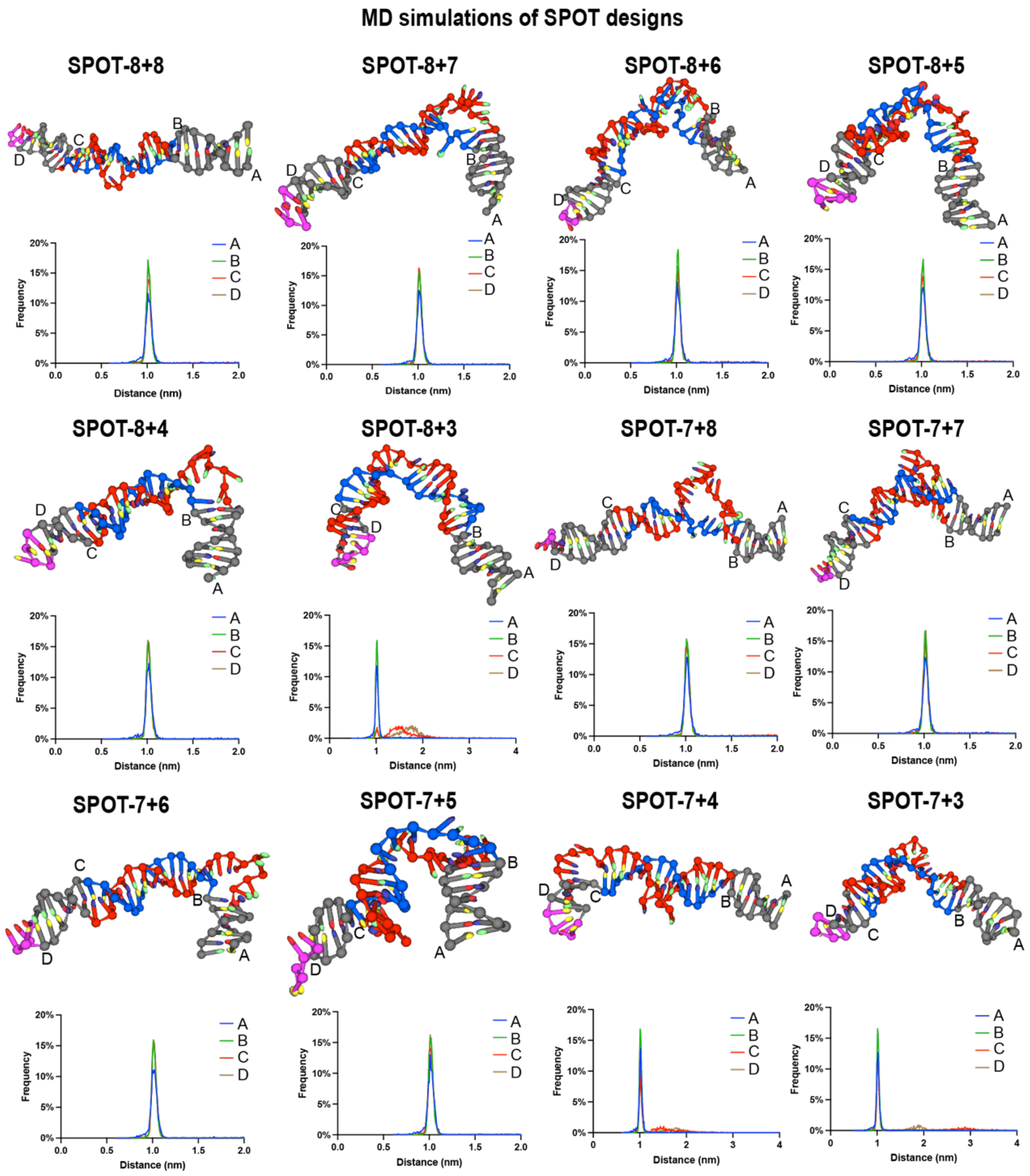

**Fig. S10 Distances between selected bases in MD models within locking domains prior to addition of target[5,6].**

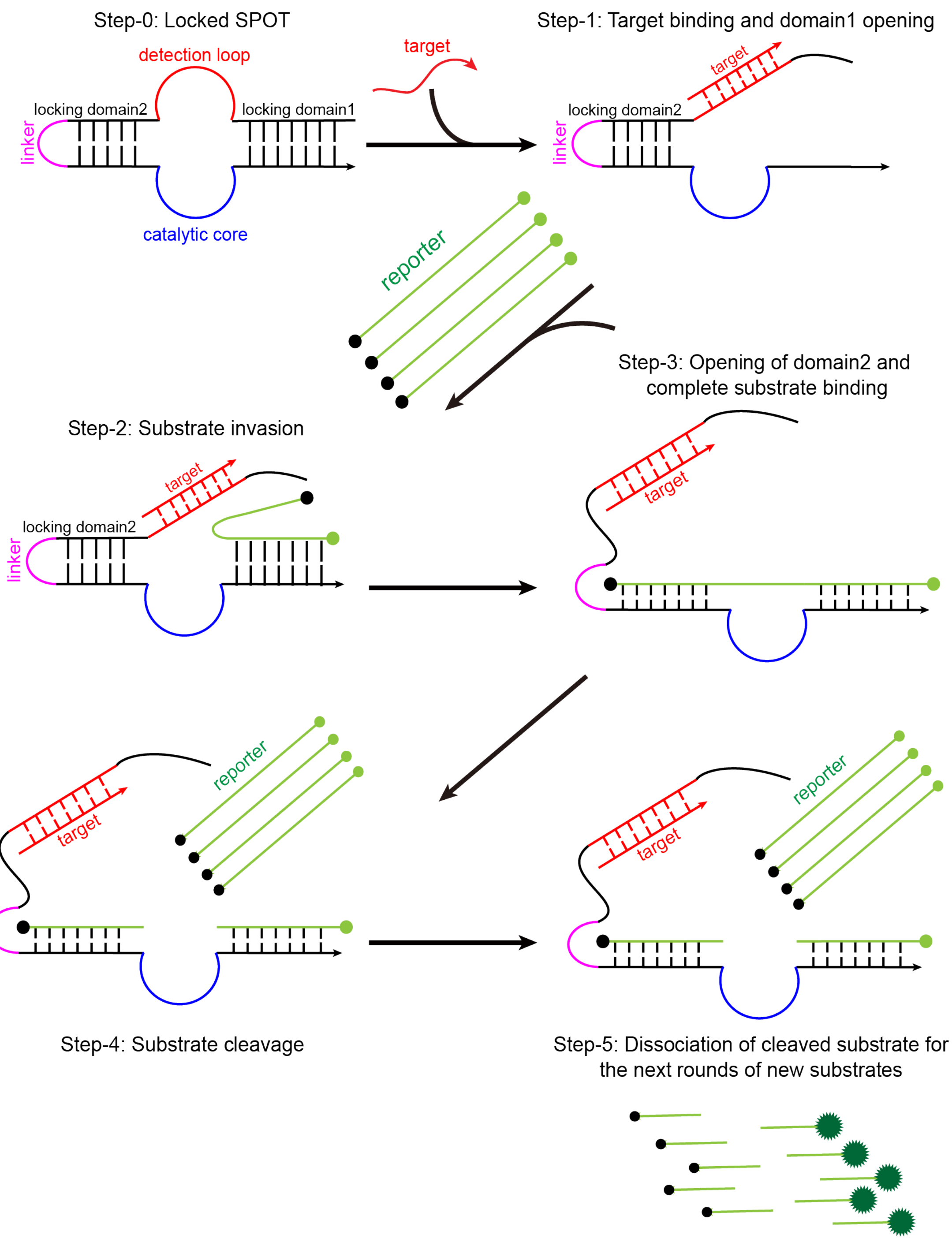

**Fig. S11 Molecular unlocking mechanism of SPOT.** domain1 is opened upon target hybridization, which leads to subsequent substrate invasion, strand displacement-induced opening of domain2, and finally cleavage of substrates.

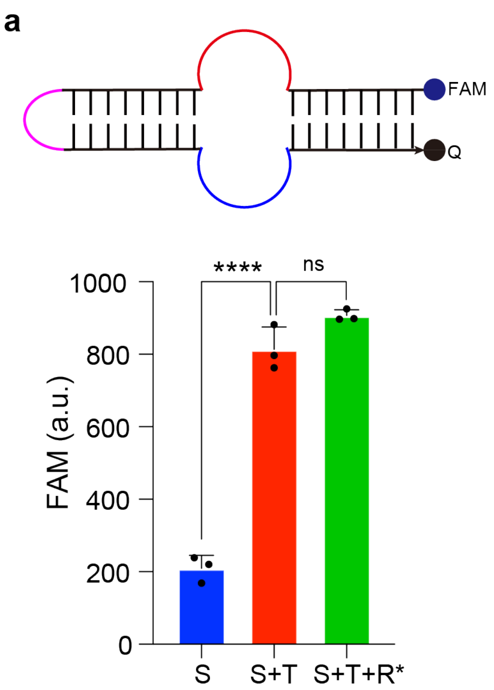

**Fig. S12 FRET experiments to investigate the molecular conformational change of SPOT-8+8 after target hybridization.** a,b, S: SPOT; T: miR-155 target; R$^*$: DNA mimic of the RNA reporter to avoid being cleaved by SPOT thus maintain it in its open state. All experimental measurements are mean ± standard deviation (SD) with n = 3.

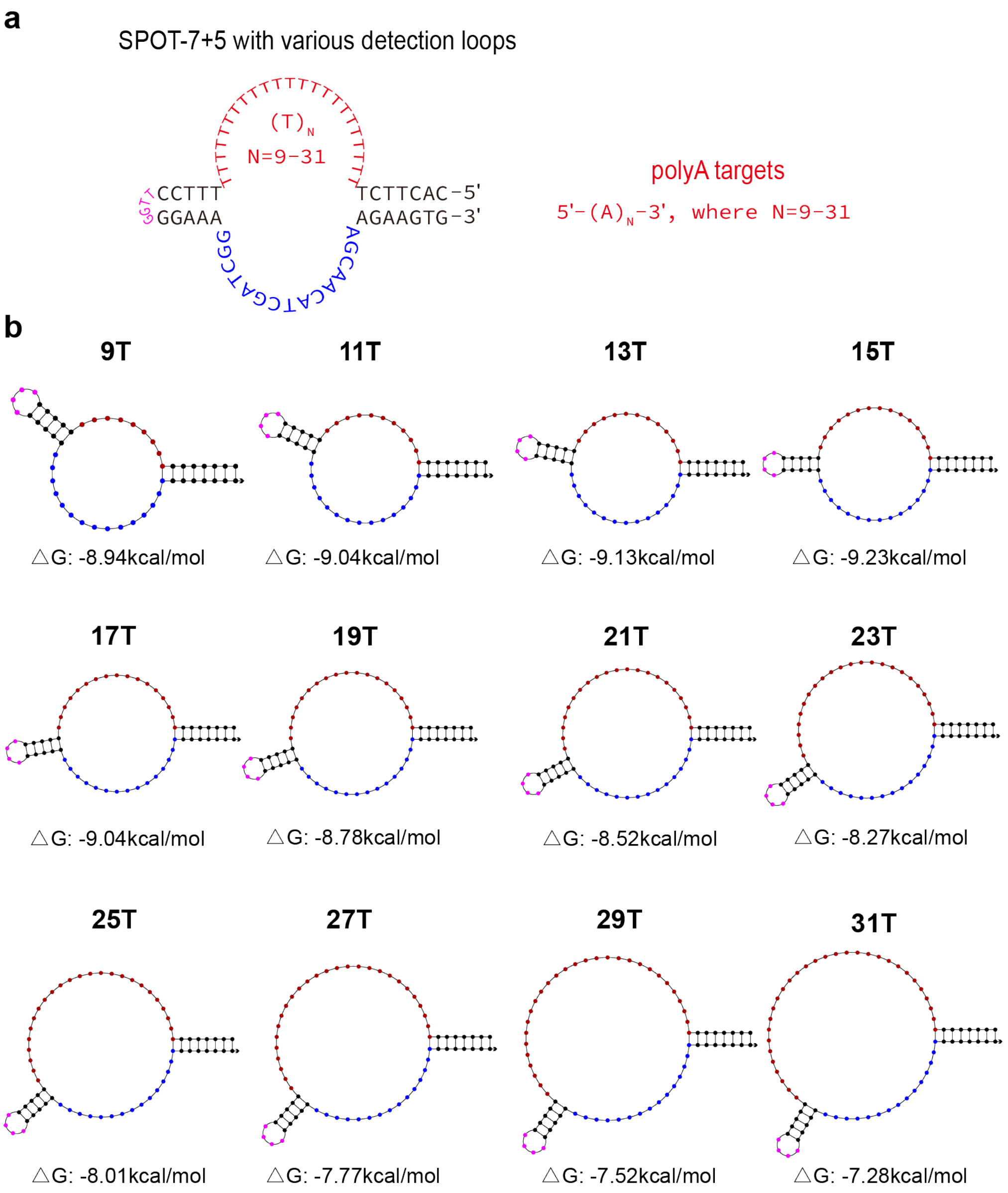

**Fig. S13 Simulation and optimization of SPOT-7+5 with various poly-T detection loop lengths (a)** Designs of SPOT-7+5 with various poly-T detection loop lengths. **(b)** Simulated secondary structures and Gibbs free energy change of SPOT-7+5 with various poly-T detection loop lengths by using NUPACK[4].

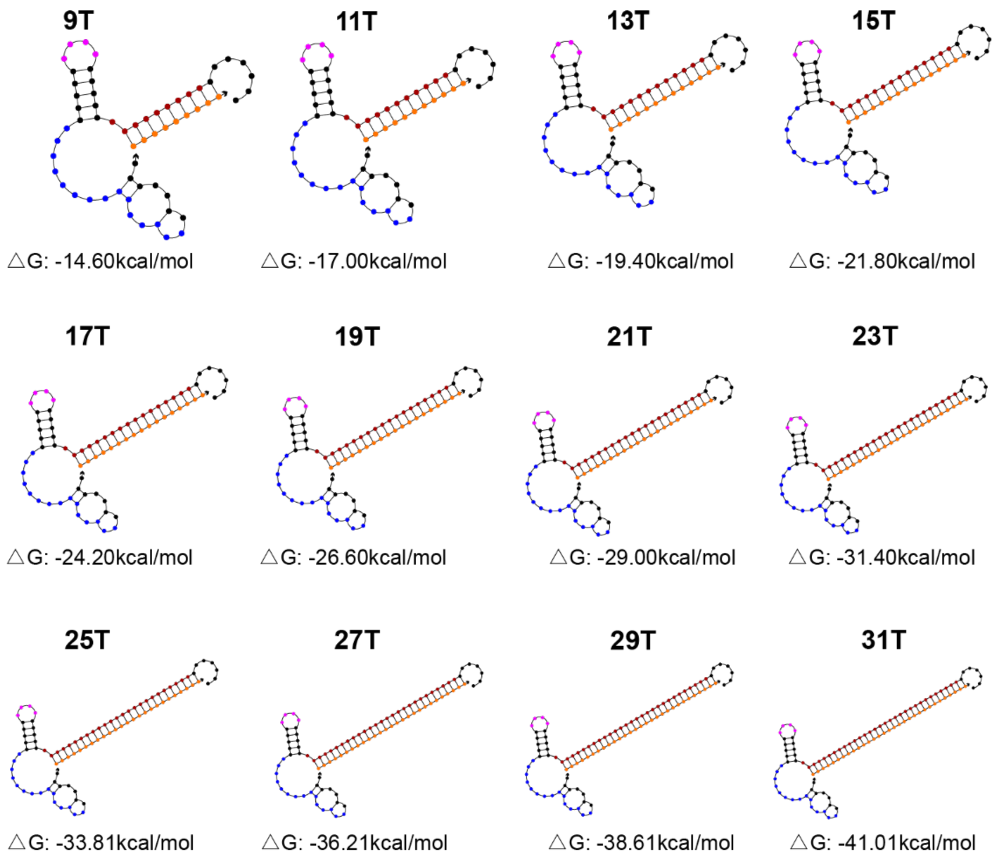

**Fig. S14 Simulated secondary structures and Gibbs free energy change of SPOT-target complexes with various poly-T detection loop lengths by using NUPACK[4].**

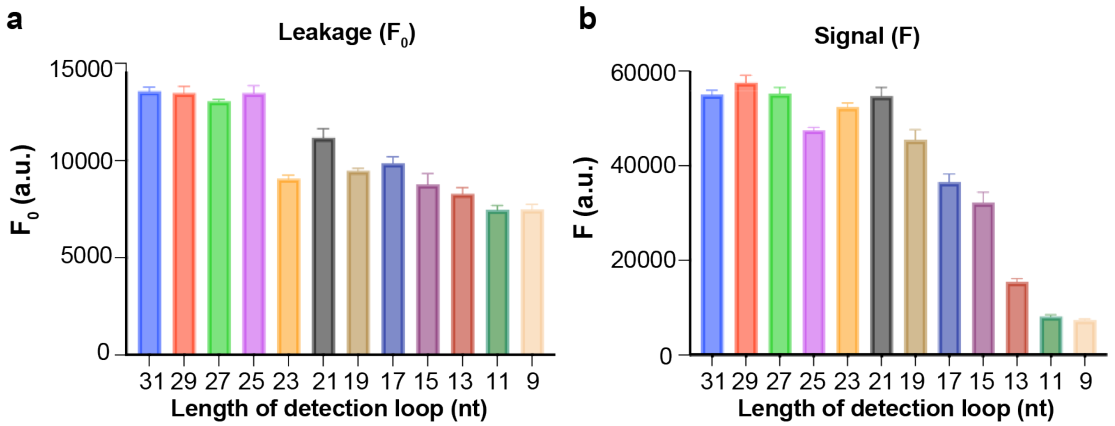

**Fig. S15 The evaluation of cleavage efficiency and accuracy for 10-23 SPOTs with different lengths of poly-T detection loops. a,** background noise ($F_0$, with target) and **b,** signal (F, without target) after 2 hours of cleavage against 10-23 reporters by all 10-23 SPOTs with various poly-T detection loop lengths. All experimental measurements are mean ± standard deviation (SD) with n = 3.

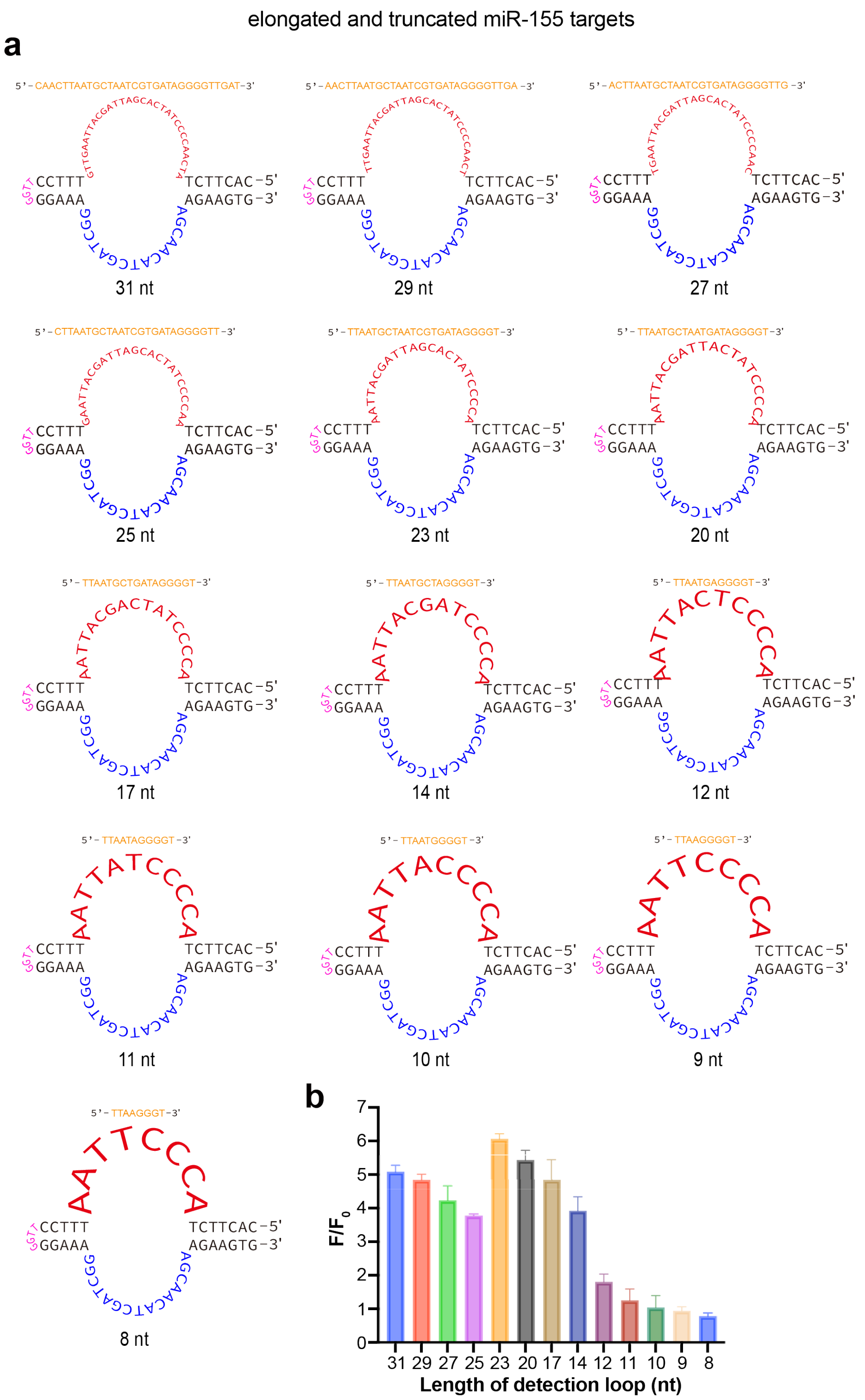

**Fig. S16 SPOT-7+5 constructed from 10-23 DNAzyme detecting elongated and truncated miR-155 targets. a,** Design of SPOT-7+5 constructed from 10-23 DNAzyme with various miR-155 detection loop lengths. **b,** Fluorescence SNR of SPOTs of various detection loops for the detection of corresponding RNA targets. All experimental measurements are mean ± standard deviation (SD) with n = 3.

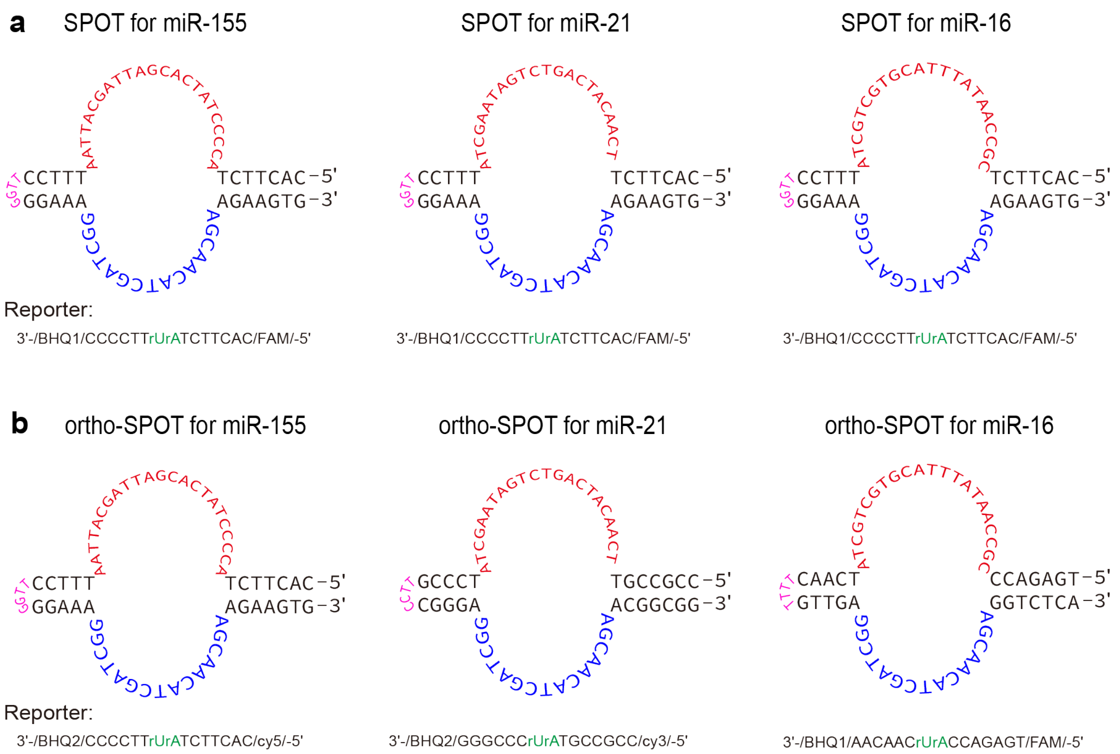

**Fig. S17 Two kinds of designs. a,** Design of SPOTs for detection of miR-155, miR-21 and miR-16. **b,** Design of the orthogonality of SPOTs for detecting multiple miRNA targets in a one-pot assay.

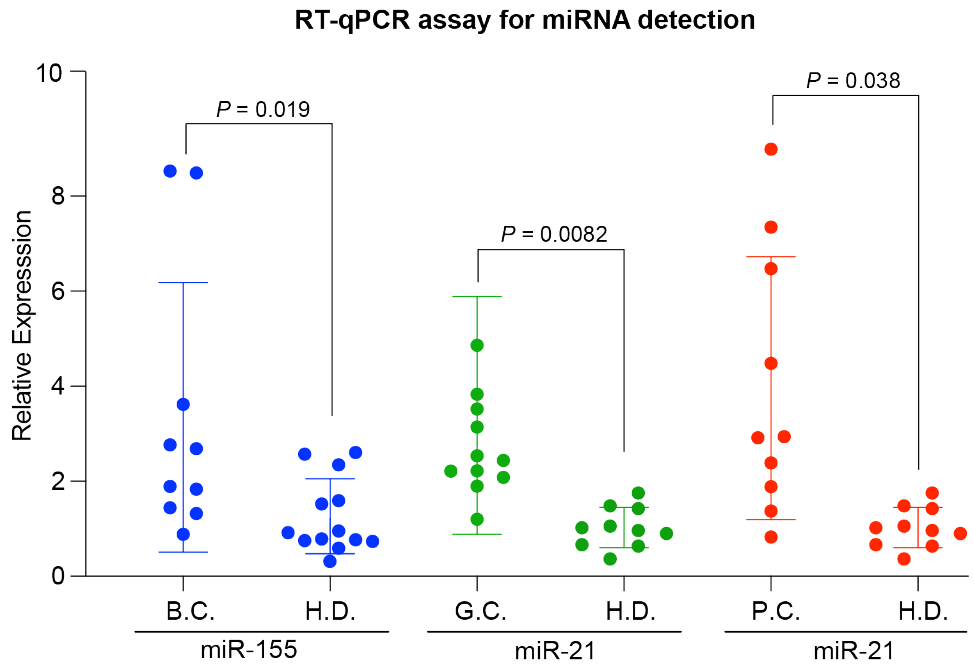

**Fig. S18 RT-qPCR for serum miRNA detection.** Scatter dot plots showing the relative expression level of miR-155 or miR-21 against miR-16 for breast cancer patients (n=10), gastric cancer patients (n=12), and prostate cancer patients (n=10), along with a number of healthy donors**.** The middle line and error bar represent the mean and SD, respectively. All experimental measurements are mean with n = 3.

### LOD linear curves for miR-155 and miR-21 based on LFA assay

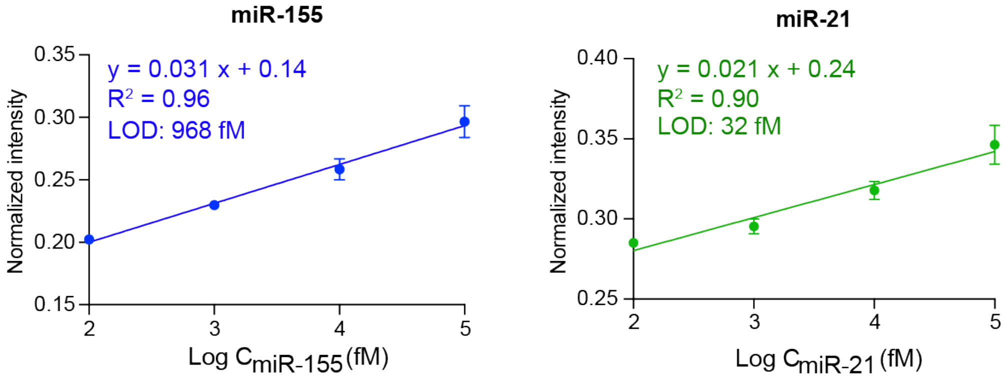

**Fig. S19 LOD linear curves for miR-155 and miR-21 based on LFA assay.** All experimental measurements are mean ± standard deviation (SD) with n = 3.

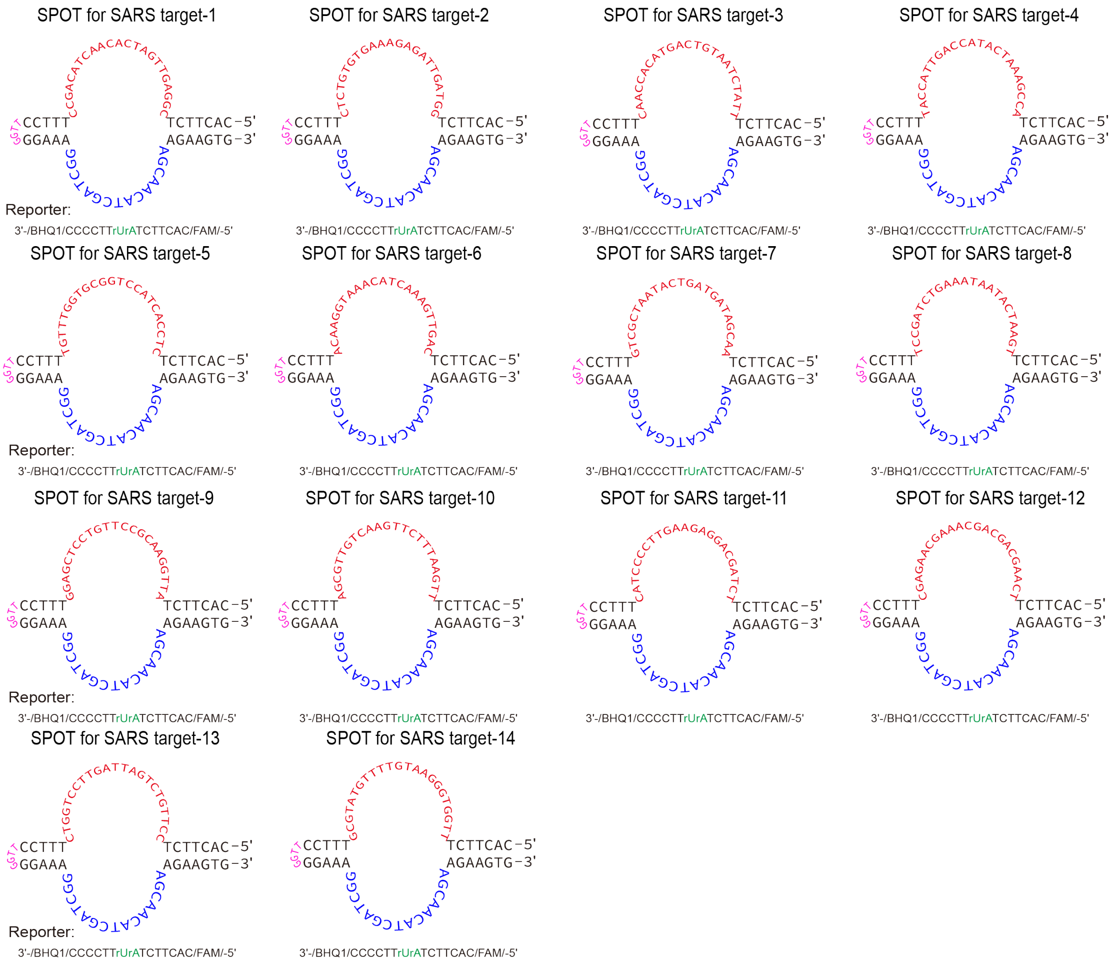

**Fig. S20 Design of SPOTs for detection of SARS-CoV-2 with different regions.**

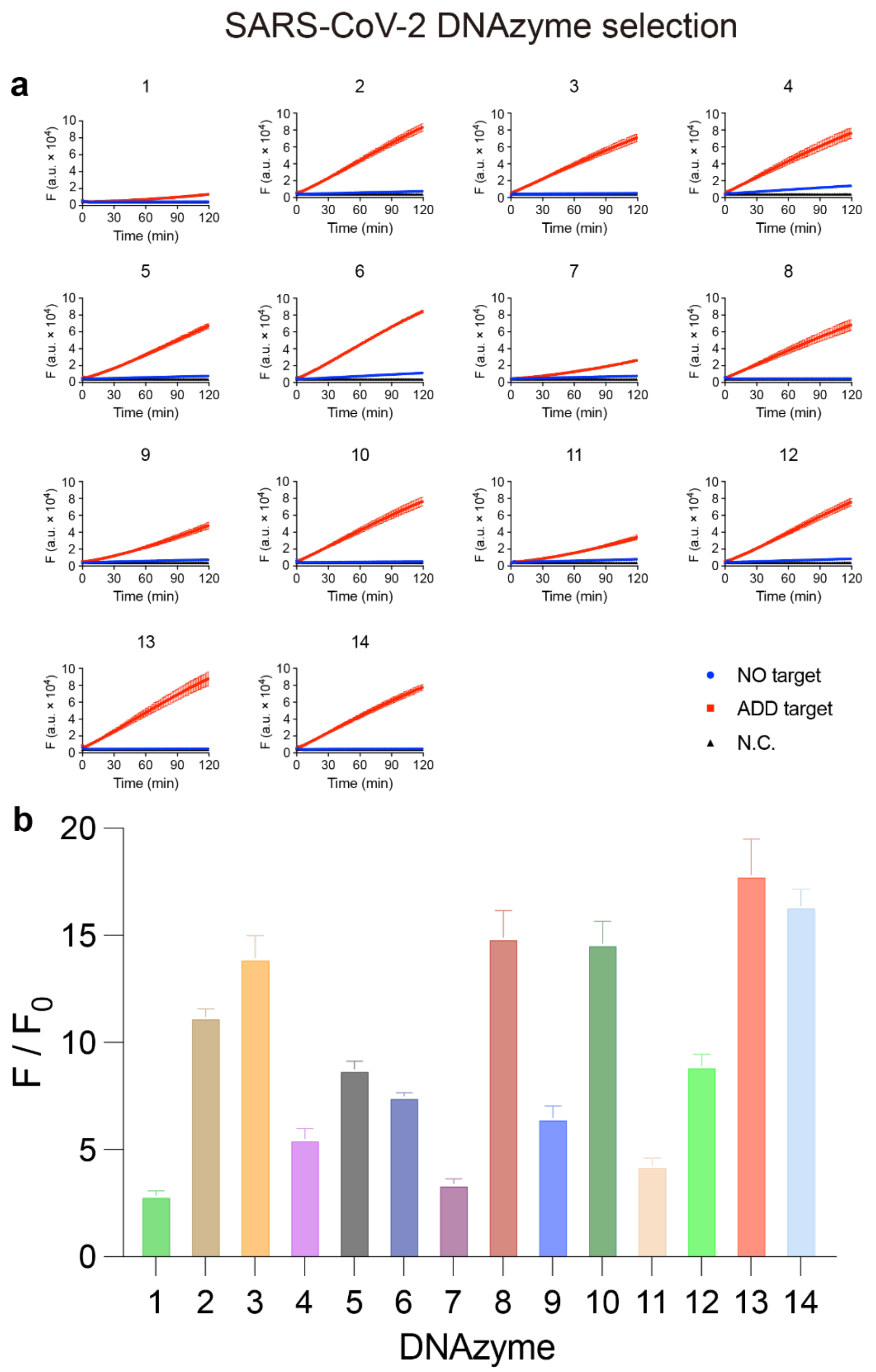

**Fig. S21 SARS-CoV-2 DNAzyme selection. a,** Kinetics for construction of SPOTs for detecting SARS-CoV-2. **b,** Fluorescence signal-to-noise ratio (SNR) of SPOTs for the detection of different regions of DNA-mimic of SARS-CoV-2 RNA after 120 mins of incubation. All experimental measurements are mean ± standard deviation (SD) with n = 3.

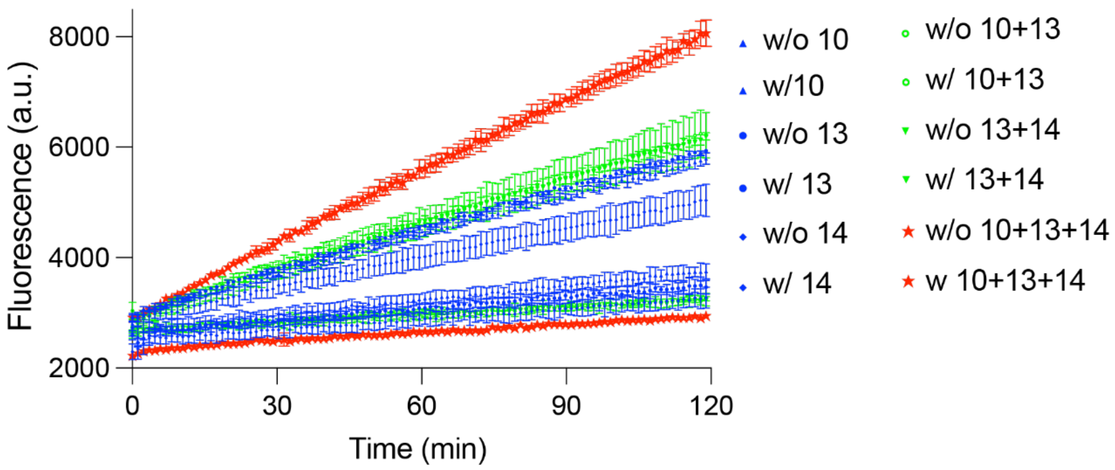

**Fig. S22 Kinetics of combinatory use of SPOT-10, SPOT-13 and SPOT-14 for SARS-CoV-2 RNA detection.** All experimental measurements are mean ± standard deviation (SD) with n = 3.

**Table S1 Oligonucleotides used in all assays.**

| No. | Sequence (from 5'-end to 3'-end) | Applied Assays |
|---|---|---|
| 10-23-SPOT-8+8 | GCACTTCTACCCCTATCACGATTAGCATTAATTTCCCCTTTTTAGGGGAAAGGCTAGCTACAACGAAGAAGTGC | Construction of SPOT |
| 10-23-SPOT-8+7 | GCACTTCTACCCCTATCACGATTAGCATTAATTTCCCCTTTTGGGGAAAGGCTAGCTACAACGAAGAAGTGC | |
| 10-23-SPOT-8+6 | GCACTTCTACCCCTATCACGATTAGCATTAATTTCCCTTTGGGGAAAGGCTAGCTACAACGAAGAAGTGC | |
| 10-23-SPOT-8+5 | GCACTTCTACCCCTATCACGATTAGCATTAATTTCCTTGGGGAAAGGCTAGCTACAACGAAGAAGTGC | |
| 10-23-SPOT-8+4 | GCACTTCTACCCCTATCACGATTAGCATTAATTTCTGGGGAAAGGCTAGCTACAACGAAGAAGTGC | |
| 10-23-SPOT-8+3 | GCACTTCTACCCCTATCACGATTAGCATTAATTTGGGGAAAGGCTAGCTACAACGAAGAAGTGC | |
| 10-23-SPOT-7+8 | CACTTCTACCCCTATCACGATTAGCATTAATTTCCCCTTTTTAGGGGAAAGGCTAGCTACAACGAAGAAGTG | |
| 10-23-SPOT-7+7 | CACTTCTACCCCTATCACGATTAGCATTAATTTCCCCTTTTGGGGAAAGGCTAGCTACAACGAAGAAGTG | |
| 10-23-SPOT-7+6 | CACTTCTACCCCTATCACGATTAGCATTAATTTCCCTTTGGGGAAAGGCTAGCTACAACGAAGAAGTG | |
| 10-23-SPOT-7+5 | CACTTCTACCCCTATCACGATTAGCATTAATTTCCTTGGGGAAAGGCTAGCTACAACGAAGAAGTG | Construction of SPOT, gel electrophoresis, miRNA detection assay, specificty assay, orthogonol assay, diagnosis of clinical samples, lateral flow assay |
| 10-23-SPOT-7+4 | CACTTCTACCCCTATCACGATTAGCATTAATTTCTGGGGAAAGGCTAGCTACAACGAAGAAGTG | Construction of SPOT |
| 10-23-SPOT-7+3 | CACTTCTACCCCTATCACGATTAGCATTAATTTGGGGAAAGGCTAGCTACAACGAAGAAGTG | |
| 10-23-SPOT-7+5-miR-21 | CACTTCTTCAACATCAGTCTGATAAGCTATTTCCTTGGGGAAAGGCTAGCTACAACGAAGAAGTG | miRNA detection assay, diagnosis of clinical samples, lateral flow assay |
| 10-23-SPOT-7+5-miR-16 | CACTTCTCGCCAATATTTACGTGCTGCTATTTCCTTGGGGAAAGGCTAGCTACAACGAAGAAGTG | miRNA detection assay, diagnosis of clinical samples |
| free-form 10-23 DNAzyme | GGGGAAAGGCTAGCTACAACGAAGAAGTG | Reporter optimization |
| reporter-RNA (linear) (FQ) (R-L) | /FAM/CACUUCUAUUUCCCC/BHQ1/ | |

| | | |
|---|---|---|
| reporter-DNA-RNA chimeric complex (MB3) (FQ) (D/R-MB3) | /FAM/CCTCACTTCTrArUTTCCCCAGG/BHQ1/ | |
| reporter-DNA-RNA chimeric complex (MB4) (FQ) (D/R-MB4) | /FAM/ACCTCACTTCTrArUTTCCCCAGGT/BHQ1/ | |
| reporter-DNA-RNA chimeric complex (linear) (FQ) (D/R-L) | /FAM/CACTTCTrArUTTCCCC/BHQ1/ | Reporter optimization, miRNA detection assay, specificty assay, orthogonol assay, diagnosis of clinical samples, SARS-CoV-2 assay |
| reporter-DNA-RNA chimeric complex (linear) | CACTTCTrArUTTCCCC | Gel electrophoresis |
| reporter-DNA-RNA chimeric complex (linear) (FAM, Biotin) | /FAM/CACTTCTrArUTTCCCC/Biotin/ | Lateral flow assay |
| 10-23-SPOT-7+5 (FQ-I) | CACTTCTACCCCTATCACGATTAGCATTAA/BHQ2 T/TTCCTTGGGGAA/cy3/AGGCTAGCTACAACGAAGAAGTG | Mechanism of SPOT |
| 10-23-SPOT-7+5 (FQ-T) | /FAM/CACTTCTACCCCTATCACGATTAGCATTAATTTCCTTGGGGAAAGGCTAGCTACAACGAAGAAGTG/BHQ1/ | |
| 10-23-SPOT-8+8 (FQ-I) | GCACTTCTACCCCTATCACGATTAGCATTAA/BHQ2 T/TTCCCCTTTTTAGGGGAA/cy3/AGGCTAGCTACAACGAAGAAGTGC | |
| 10-23-SPOT-8+8 (FQ-T) | /FAM/GCACTTCTACCCCTATCACGATTAGCATTAATTTCCCCTTTTTAGGGGAAAGGCTAGCTACAACGAAGAAGTGC/BHQ1/ | |
| DNA mimic of DNA-RNA chimeric complex reporter | CACTTCTATTTCCCC | |
| SPOT-7+5-31T | CACTTCTTTTTTTTTTTTTTTTTTTTTTTTTTTTTTTTTTTCCTTGGGGAAAGGCTAGCTACAACGAAGAAGTG | SPOT mechanism |
| SPOT-7+5-29T | CACTTCTTTTTTTTTTTTTTTTTTTTTTTTTTTTTTTTTCCTTGGGGAAAGGCTAGCTACAACGAAGAAGTG | |
| SPOT-7+5-27T | CACTTCTTTTTTTTTTTTTTTTTTTTTTTTTTTTTTTCCTTGGGGAAAGGCTAGCTACAACGAAGAAGTG | |
| SPOT-7+5-25T | CACTTCTTTTTTTTTTTTTTTTTTTTTTTTTTTTTCCTTGGGGAAAGGCTAGCTACAACGAAGAAGTG | |
| SPOT-7+5-23T | CACTTCTTTTTTTTTTTTTTTTTTTTTTTTTTTCCTTGGGGAAAGGCTAGCTACAACGAAGAAGTG | |

| | | |
|---|---|---|
| SPOT-7+5-21T | CACTTCTTTTTTTTTTTTTTTTTTTTTTTTTCCTTGGGGAAAGGCTAGCTACAACGAAGAAGTG | |
| SPOT-7+5-19T | CACTTCTTTTTTTTTTTTTTTTTTTTTTTCCTTGGGGAAAGGCTAGCTACAACGAAGAAGTG | |
| SPOT-7+5-17T | CACTTCTTTTTTTTTTTTTTTTTTTTTCCTTGGGGAAAGGCTAGCTACAACGAAGAAGTG | |
| SPOT-7+5-15T | CACTTCTTTTTTTTTTTTTTTTTTTCCTTGGGGAAAGGCTAGCTACAACGAAGAAGTG | |
| SPOT-7+5-13T | CACTTCTTTTTTTTTTTTTTTTTCCTTGGGGAAAGGCTAGCTACAACGAAGAAGTG | |
| SPOT-7+5-11T | CACTTCTTTTTTTTTTTTTTTCCTTGGGGAAAGGCTAGCTACAACGAAGAAGTG | |
| SPOT-7+5-9T | CACTTCTTTTTTTTTTTTTCCTTGGGGAAAGGCTAGCTACAACGAAGAAGTG | |
| target-31A | AAAAAAAAAAAAAAAAAAAAAAAAAAAAAAA | |
| target-29A | AAAAAAAAAAAAAAAAAAAAAAAAAAAAA | |
| target-27A | AAAAAAAAAAAAAAAAAAAAAAAAAAA | |
| target-25A | AAAAAAAAAAAAAAAAAAAAAAAAA | |
| target-23A | AAAAAAAAAAAAAAAAAAAAAAA | |
| target-21A | AAAAAAAAAAAAAAAAAAAAA | |
| target-19A | AAAAAAAAAAAAAAAAAAA | |
| target-17A | AAAAAAAAAAAAAAAAA | |
| target-15A | AAAAAAAAAAAAAAA | |
| target-13A | AAAAAAAAAAAAA | |
| target-11A | AAAAAAAAAAA | |
| target-9A | AAAAAAAAA | |
| SPOT-7+5-155-31nt | CACTTCTATCAACCCCTATCACGATTAGCATTAAGTTGTTTCCTTGGGGAAAGGCTAGCTACAACGAAGAAGTG | SPOT mechanism |
| SPOT-7+5-155-29nt | CACTTCTTCAACCCCTATCACGATTAGCATTAAGTTTTTCCTTGGGGAAAGGCTAGCTACAACGAAGAAGTG | |
| SPOT-7+5-155-27nt | CACTTCTCAACCCCTATCACGATTAGCATTAAGTTTTCCTTGGGGAAAGGCTAGCTACAACGAAGAAGTG | |

| | |
|---|---|
| SPOT-7+5-155-25nt | CACTTCTAACCCCTATCACGATTAGCATTAAGTTTCCTTGGGGAAAGGCTAGCTACAACGAAGAAGTG |
| SPOT-7+5-155-23nt | CACTTCTACCCCTATCACGATTAGCATTAATTTCCTTGGGGAAAGGCTAGCTACAACGAAGAAGTG |
| SPOT-7+5-155-20nt | CACTTCTACCCCTATCATTAGCATTAATTTCCTTGGGGAAAGGCTAGCTACAACGAAGAAGTG |
| SPOT-7+5-155-17nt | CACTTCTACCCCTATCAGCATTAATTTCCTTGGGGAAAGGCTAGCTACAACGAAGAAGTG |
| SPOT-7+5-155-14nt | CACTTCTACCCCTAGCATTAATTTCCTTGGGGAAAGGCTAGCTACAACGAAGAAGTG |
| SPOT-7+5-155-12nt | CACTTCTACCCCTCATTAATTTCCTTGGGGAAAGGCTAGCTACAACGAAGAAGTG |
| SPOT-7+5-155-11nt | CACTTCTACCCCTATTAATTTCCTTGGGGAAAGGCTAGCTACAACGAAGAAGTG |
| SPOT-7+5-155-10nt | CACTTCTACCCCATTAATTTCCTTGGGGAAAGGCTAGCTACAACGAAGAAGTG |
| SPOT-7+5-155-9nt | CACTTCTACCCCTTAATTTCCTTGGGGAAAGGCTAGCTACAACGAAGAAGTG |
| SPOT-7+5-155-8nt | CACTTCTACCCTTAATTTCCTTGGGGAAAGGCTAGCTACAACGAAGAAGTG |
| DNA mimic-miR-155-31nt | CAACTTAATGCTAATCGTGATAGGGGTTGAT |
| DNA mimic-miR-155-29nt | AACTTAATGCTAATCGTGATAGGGGTTGA |
| DNA mimic-miR-155-27nt | ACTTAATGCTAATCGTGATAGGGGTTG |
| DNA mimic-miR-155-25nt | CTTAATGCTAATCGTGATAGGGGTT |
| DNA mimic-miR-155-23nt | TTAATGCTAATCGTGATAGGGGT |
| DNA mimic-miR-155-20nt | TTAATGCTAATGATAGGGGT |
| DNA mimic-miR-155-17nt | TTAATGCTGATAGGGGT |
| DNA mimic-miR-155-14nt | TTAATGCTAGGGGT |
| DNA mimic-miR-155-12nt | TTAATGAGGGGT |
| DNA mimic-miR-155-11nt | TTAATAGGGGT |
| DNA mimic-miR-155-10nt | TTAATGGGGT |
| DNA mimic-miR-155-9nt | TTAAGGGGT |

| | | |
|---|---|---|
| DNA mimic-miR-155-8nt | TTAAGGGT | |
| miR-155 | UUAAUGCUAAUCGUGAUAGGGGU | Construction of SPOT, mechanism of SPOT, miRNA detection assay, specificity assay, gel electrophoresis, lateral flow assay |
| miR-21 | UAGCUUAUCAGACUGAUGUUGA | miRNA detection assay, specificity assay, lateral flow assay |
| miR-103a | AGCAGCAUUGUACAGGGCUAUGA | specificity assay |
| miR-29c | UGACCGAUUUCUCCUGGUGUUC | |
| miR-16 | UAGCAGCACGUAAAUAUUGGCG | miRNA detection assay, specificity assay |
| ortho-SPOT-7+5-16 | TGAGACCCGCCAATATTTACGTGCTGCTATCAACTTTTGTTGAGGCTAGCTACAACGAGGTCTCA | Orthogonal assay |
| ortho-SPOT-7+5-21 | CCGCCGTTCAACATCAGTCTGATAAGCTATCCCGTTCCCGGGAGGCTAGCTACAACGAACGGCGG | |
| ortho-reporter-16 (FAM) | /FAM/TGAGACCrArUCAACAA/BHQ1/ | |
| ortho-reporter-21 (cy3) | /cy3/CCGCCGTrArUCCCGGG/BHQ2/ | |
| ortho-reporter-155 (cy5) | /cy5/CACTTCTrArUTTCCCC/BHQ2/ | |
| miR-155 SNV-5 | UUAAGGCUAAUCGUGAUAGGGGU | Specificity assay |
| miR-155 SNV-7 | UUAAUGAUAAUCGUGAUAGGGGU | |
| miR-155 SNV-9 | UUAAUGCUGAUCGUGAUAGGGGU | |
| miR-155 SNV-11 | UUAAUGCUAACCGUGAUAGGGGU | |
| miR-155 SNV-17 | UUAAUGCUAAUCGUGACAGGGGU | |
| miR-155 SNV-19 | UUAAUGCUAAUCGUGAUAUGGGU | |
| miR-155 SNV-21 | UUAAUGCUAAUCGUGAUAGGCGU | |
| miR-155 scramble | GAUGGUUACGAUUAUUAGGCUAG | |
| 13PD1-SPOT-11+5+4 | GATCTCCTCCTACCCCTATCACGATTAGCATTAAGCTTCAGAGGAAGCTATACCGGGCAACTATTGCCTCGTCA | 13PD1 assay |

| | | |
|---|---|---|
| | TCGCTATTTTCTGCGAGGAGGAGATC | |
| 13PD1- SPOT-12+5+4 | AGATCTCCTCCTACCCCTATCACGATTAGCATTAAGCTTCAGAGGAAGCTATACCGGGCAACTATTGCCTCGTC ATCGCTATTTTCTGCGAGGAGGAGATCT | |
| 13PD1- SPOT-13+5+4 | CAGATCTCCTCCTACCCCTATCACGATTAGCATTAAGCTTCAGAGGAAGCTATACCGGGCAACTATTGCCTCGT CATCGCTATTTTCTGCGAGGAGGAGATCTG | |
| 13PD1- SPOT-14+5+4 | CCAGATCTCCTCCTACCCCTATCACGATTAGCATTAAGCTTCAGAGGAAGCTATACCGGGCAACTATTGCCTCG TCATCGCTATTTTCTGCGAGGAGGAGATCTGG | |
| 13PD1- SPOT-12+4+5 | AGATCTCCTCCTACCCCTATCACGATTAGCATTAAGCTTAGAGGAAGCTATACCGGGCAACTATTGCCTCGTCAT CGCTATTTTCTGCGAGGAGGAGATCT | |
| 13PD1- SPOT-13+4+5 | CAGATCTCCTCCTACCCCTATCACGATTAGCATTAAGCTTAGAGGAAGCTATACCGGGCAACTATTGCCTCGTC ATCGCTATTTTCTGCGAGGAGGAGATCTG | |
| 13PD1- SPOT-14+4+5 | CCAGATCTCCTCCTACCCCTATCACGATTAGCATTAAGCTTAGAGGAAGCTATACCGGGCAACTATTGCCTCGT CATCGCTATTTTCTGCGAGGAGGAGATCTGG | |
| freeform 13PD1 DNAzyme | AGAGGAAGCTATACCGGGCAACTATTGCCTCGTCATCGCTATTTTCTGCGAGGAGGAGA | |
| 13PD1-reporter (FQ) | /FAM/TCTCCTCCTACTGCTGCTTCCTCT/BHQ1/ | |
| DNA-mimic miR-155 | TTAATGCTAATCGTGATAGGGGT | |
| target 1-SARS-CoV-2 | GGCTGTAGTTGTGATCAACTCCG | SARS-CoV-2 assay |
| target 2-SARS-CoV-2 | GAGACACACTTTCTCTAACTACC | |
| target 3-SARS-CoV-2 | GTTGGTGTACTGACATTAGATAA | |
| target 4-SARS-CoV-2 | ATGGTAACTGGTATGATTTCGGT | |
| target 5-SARS-CoV-2 | ACAAACCACGCCAGGTAGTGGAG | |
| target 6-SARS-CoV-2 | TGTTCCATTTGTAGTTTCAACTG | |
| target 7-SARS-CoV-2 | CAGCGATTATGACTACTATCGTT | |
| target 8-SARS-CoV-2 | AGGCTAGACTTTATTATGATTCA | |

| | |
|---|---|
| target 9-SARS-CoV-2 | CCTCGAGGACAAGGCGTTCCAAT |
| target 10-SARS-CoV-2 | TCGCAACAGTTCAAGAAATTCAA |
| target 11-SARS-CoV-2 | GTAGGGGAACTTCTCCTGCTAGA |
| target 12-SARS-CoV-2 | GCTCTTGCTTTGCTGCTGCTTGA |
| target 13-SARS-CoV-2 | GACCAGGAACTAATCAGACAAGG |
| target 14-SARS-CoV-2 | CGCATACAAAACATTCCCACCAA |
| RNA-target 3-SARS-CoV-2 | GUUGGUGUACUGACAUUAGAUAA |
| RNA-target 8-SARS-CoV-2 | AGGCUAGACUUUAUUAUGAUUCA |
| RNA-target 10-SARS-CoV-2 | UCGCAACAGUUCAAGAAAUUCAA |
| RNA-target 13-SARS-CoV-2 | GACCAGGAACUAAUCAGACAAGG |
| RNA-target 14-SARS-CoV-2 | CGCAUACAAAACAUUCCCACCAA |
| SPOT-target 1-SARS-CoV-2 | CACTTCTCGGAGTTGATCACAACTACAGCCTTTCCTTGGGGAAAGGCTAGCTACAACGAAGAAGTG |
| SPOT-target 2-SARS-CoV-2 | CACTTCTGGTAGTTAGAGAAAGTGTGTCTCTTTCCTTGGGGAAAGGCTAGCTACAACGAAGAAGTG |
| SPOT-target 3-SARS-CoV-2 | CACTTCTTTATCTAATGTCAGTACACCAACTTTCCTTGGGGAAAGGCTAGCTACAACGAAGAAGTG |
| SPOT-target 4-SARS-CoV-2 | CACTTCTACCGAAATCATACCAGTTACCATTTTCCTTGGGGAAAGGCTAGCTACAACGAAGAAGTG |
| SPOT-target 5-SARS-CoV-2 | CACTTCTCTCCACTACCTGGCGTGGTTTGTTTTCCTTGGGGAAAGGCTAGCTACAACGAAGAAGTG |
| SPOT-target 6-SARS-CoV-2 | CACTTCTCAGTTGAAACTACAAATGGAACATTTCCTTGGGGAAAGGCTAGCTACAACGAAGAAGTG |
| SPOT-target 7-SARS-CoV-2 | CACTTCTAACGATAGTAGTCATAATCGCTGTTTCCTTGGGGAAAGGCTAGCTACAACGAAGAAGTG |
| SPOT-target 8-SARS-CoV-2 | CACTTCTTGAATCATAATAAAGTCTAGCCTTTTCCTTGGGGAAAGGCTAGCTACAACGAAGAAGTG |
| SPOT-target 9-SARS-CoV-2 | CACTTCTATTGGAACGCCTTGTCCTCGAGGTTTCCTTGGGGAAAGGCTAGCTACAACGAAGAAGTG |
| SPOT-target 10-SARS-CoV-2 | CACTTCTTTGAATTTCTTGAACTGTTGCGATTTCCTTGGGGAAAGGCTAGCTACAACGAAGAAGTG |
| SPOT-target 11-SARS-CoV-2 | CACTTCTTCTAGCAGGAGAAGTTCCCCTACTTTCCTTGGGGAAAGGCTAGCTACAACGAAGAAGTG |

| | |
|---|---|
| SPOT-target 12-SARS-CoV-2 | CACTTCTTCAAGCAGCAGCAAAGCAAGAGCTTTCCTTGGGGAAAGGCTAGCTACAACGAAGAAGTG |
| SPOT-target 13-SARS-CoV-2 | CACTTCTCCTTGTCTGATTAGTTCCTGGTCTTTCCTTGGGGAAAGGCTAGCTACAACGAAGAAGTG |
| SPOT-target 14-SARS-CoV-2 | CACTTCTTTGGTGGGAATGTTTTGTATGCGTTTCCTTGGGGAAAGGCTAGCTACAACGAAGAAGTG |

**Table S2 Information of clinical samples.**

| Breast Cancer-Fluorescence | |
|---|---|
| No. | Clinical diagnose |
| B-1 | Breast malignancy |
| B-2 | Breast malignancy |
| B-3 | Breast malignancy |
| B-4 | Breast malignancy |
| B-5 | Breast malignancy |
| B-6 | Breast malignancy |
| B-7 | Breast malignancy |
| B-8 | Breast malignancy |
| B-9 | Breast malignancy |
| B-10 | Breast malignancy |
| Gastric Cancer-Fluorescence | |
| No. | Clinical diagnose |
| G-1 | Gastric malignant tumor |
| G-2 | Gastric malignant tumor |
| G-3 | Gastric malignant tumor |
| G-4 | Gastric malignant tumor |
| G-5 | Gastric malignant tumor |
| G-6 | Gastric malignant tumor |
| G-7 | Gastric malignant tumor |
| G-8 | Gastric malignant tumor |
| G-9 | Gastric malignant tumor |
| G-10 | Gastric malignant tumor |
| G-11 | Gastric malignant tumor |
| G-12 | Gastric malignant tumor |
| Prostate Cancer-Fluorescence | |
| No. | Clinical diagnose |
| P-1 | Prostate Cancer |
| P-2 | Prostate Cancer |
| P-3 | Prostate Cancer |
| P-4 | Prostate Cancer |
| P-5 | Prostate Cancer |
| P-6 | Prostate Cancer |
| P-7 | Prostate Cancer |
| P-8 | Prostate Cancer |
| P-9 | Prostate Cancer |
| P-10 | Prostate Cancer |
| Healthy donors-Fluorescence | |
| No. | Clinical diagnose |
| C-1 | (-) |
| C-2 | (-) |

| | |
|---|---|
| C-3 | (-) |
| C-4 | (-) |
| C-5 | (-) |
| C-6 | (-) |
| C-7 | (-) |
| C-8 | (-) |
| C-9 | (-) |
| C-10 | (-) |
| C-11 | (-) |
| C-12 | (-) |
| C-13 | (-) |
| Breast Cancer-LFA | |
| No. | Clinical diagnose |
| B-11 | Breast malignancy |
| B-12 | Breast malignancy |
| B-13 | Breast malignancy |
| B-14 | Breast malignancy |
| B-15 | Breast malignancy |
| B-16 | Breast malignancy |
| Gastric Cancer-LFA | |
| No. | Clinical diagnose |
| G-13 | Gastric malignant tumor |
| G-14 | Gastric malignant tumor |
| G-15 | Gastric malignant tumor |
| G-16 | Gastric malignant tumor |
| Prostate Cancer-LFA | |
| No. | Clinical diagnose |
| P-11 | Prostate Cancer |
| P-12 | Prostate Cancer |
| P-13 | Prostate Cancer |
| P-14 | Prostate Cancer |
| P-15 | Prostate Cancer |
| P-16 | Prostate Cancer |
| Healthy donors-LFA (compared with Breast Cancer-LFA) | |
| No. | Clinical diagnose |
| C-14 | (-) |
| C-15 | (-) |
| C-16 | (-) |
| C-17 | (-) |
| C-18 | (-) |
| C-19 | (-) |
| Healthy donors-LFA (compared with Gastric Cancer-LFA) | |

| No. | Clinical diagnose |
|---|---|
| C-20 | (-) |
| C-21 | (-) |
| C-22 | (-) |
| C-23 | (-) |
| Healthy donors-LFA (compared with Prostate Cancer-LFA) | |
| No. | Clinical diagnose |
| C-24 | (-) |
| C-25 | (-) |
| C-26 | (-) |
| C-27 | (-) |
| C-28 | (-) |
| C-29 | (-) |

| SARS-CoV-2-Fluorescence | |
|---|---|
| | Average CT |
| Po-1 | 21-22 |
| Po-2 | 24-25 |
| Po-3 | 23-24 |
| Po-4 | 32-33 |
| Po-5 | 28-29 |
| Po-6 | 27-28 |
| Po-7 | 29-30 |
| Po-8 | 27-28 |
| Ng-1 | (-) |
| Ng-2 | (-) |
| Ng-3 | (-) |
| Ng-4 | (-) |
| Ng-5 | (-) |
| Ng-6 | (-) |
| Ng-7 | (-) |
| Ng-8 | (-) |
| SARS-CoV-2-LFA | |
| | Average CT |
| Po-1 | 19-20 |
| Po-2 | 21-22 |
| Po-3 | 20-21 |
| Po-4 | 22-23 |
| Po-5 | 20-21 |
| Po-6 | 19-20 |
| Ng-1 | (-) |
| Ng-2 | (-) |
| Ng-3 | (-) |

| | |
|---|---|
| Ng-4 | (-) |

**Table S3 Comparison of SPOT with other exemplary miRNA and virus detection methods.**

| method | major components (number of enzyme and probes) | DNAzyme / CRISPR strategy | pre-amplification | one-pot reaction (steps) | LOD | assay time | cost per test |
|---|---|---|---|---|---|---|---|
| SPOT | 0 enzyme, 2 probes | loop activated DNAzyme | No | Yes | miRNAs: femtomolar; SARS-CoV-2 RNA: attomolar | 2.25 h | $0.02 |
| EXTRA-CRISPR[14] | 3 enzymes, 3 probes | cis- and trans-cleavage of CRISPR-Cas12a | Yes, RCA | Yes | miRNAs: femtomolar | 1.5 h | $0.60 |
| DNAzyme motor[9] | 0 enzyme, 3 probes, AuNP | toehold activated DNAzyme | No | Yes | miRNAs: picomolar | 1.33h | NA |
| REVEALR[10] | 3 enzymes, 5 probes, FANA | split-and-resume DNAzyme | Yes, RPA | No, 2 steps | SARS-CoV-2 RNA: attomolar | 2.5 h | NA |
| DNAzyme-MB[15] | 0 enzyme, 2 probes | toehold activated DNAzyme | No | Yes | DNA target: picomolar | 3 h | NA |
| SHERLOCK[16] | 4 enzymes, 4 probes | trans-cleavage of CRISPR-Cas13a | Yes, RPA | No, 2 steps | ZIKA RNA: attomolar | 2.5-5 h | $0.61 |
| MNAzymes[11] | 0 enzyme, 3 probes | split-and-resume DNAzyme | No | Yes | DNA target: picomolar | 2h | NA |
| CRISPR-HCR[17] | 1 enzyme, 5 probes | HCR and trans-cleavage of CRISPR-Cas12a | Yes | No, 2 steps | miRNAs: femtomolar | 50 min | NA |
